# Tauroursodeoxycholic Acid (TUDCA) Reduces ER Stress and Lessens Disease Activity in Ulcerative Colitis

**DOI:** 10.1101/2025.04.02.25322684

**Authors:** Vladimir Lamm, Katherine Huang, Ruishu Deng, Siyan Cao, Miao Wang, Saeed Soleymanjahi, Thanyarat Promlek, Rachel Rodgers, Deanna Davis, Darren Nix, Guadalupe Oliva Escudero, Yan Xie, Chien-Huan Chen, Anas Gremida, Richard P Rood, Ta-Chiang Liu, Megan T Baldridge, Parakkal Deepak, Nicholas O Davidson, Randal J Kaufman, Matthew A Ciorba

## Abstract

**Background and Aims:** In inflammatory bowel disease, protein misfolding in the endoplasmic reticulum (ER) potentiates epithelial barrier dysfunction and impairs mucosal healing. Tauroursodeoxycholic acid (TUDCA), a naturally occurring bile acid, acts as a chemical chaperone to reduce protein aggregation and colitis severity in preclinical models. We conducted an open label trial evaluating oral TUDCA as therapy in patients with active ulcerative colitis (UC).

**Methods:** Patients with moderate-to-severely active UC (Mayo score ≥6, endoscopic subscore ≥1) received oral TUDCA at 1.75 or 2 g/day for 6 weeks. Exclusion criteria included known hepatic disorders or change in UC therapy within 60 days. Clinical disease activity questionnaires, endoscopy with biopsy, blood, and stool were collected at enrollment and after 6 weeks. The primary outcome measure was change in ER stress markers while safety, tolerability and change in UC disease activity were secondary outcomes.

**Results:** Thirteen participants completed the study with eleven evaluable for clinical response. TUDCA was well-tolerated with transient dyspepsia being the most common side effect. Mucosal biopsies revealed significant reductions in ER stress and inflammation as well as an increase in markers of epithelial restitution. Clinical, endoscopic, and histologic disease activity were significantly improved at week 6 (mean total Mayo Score: 9 to 4.5, p<0.001).

**Conclusions:** Six weeks of oral TUDCA treatment was well-tolerated in patients with active ulcerative colitis and promoted mucosal healing, lessened ER stress, and reduced clinical disease activity. A randomized controlled trial of adjunctive TUDCA therapy in patients with UC is warranted.

**GRAPHICAL ABSTRACT:** 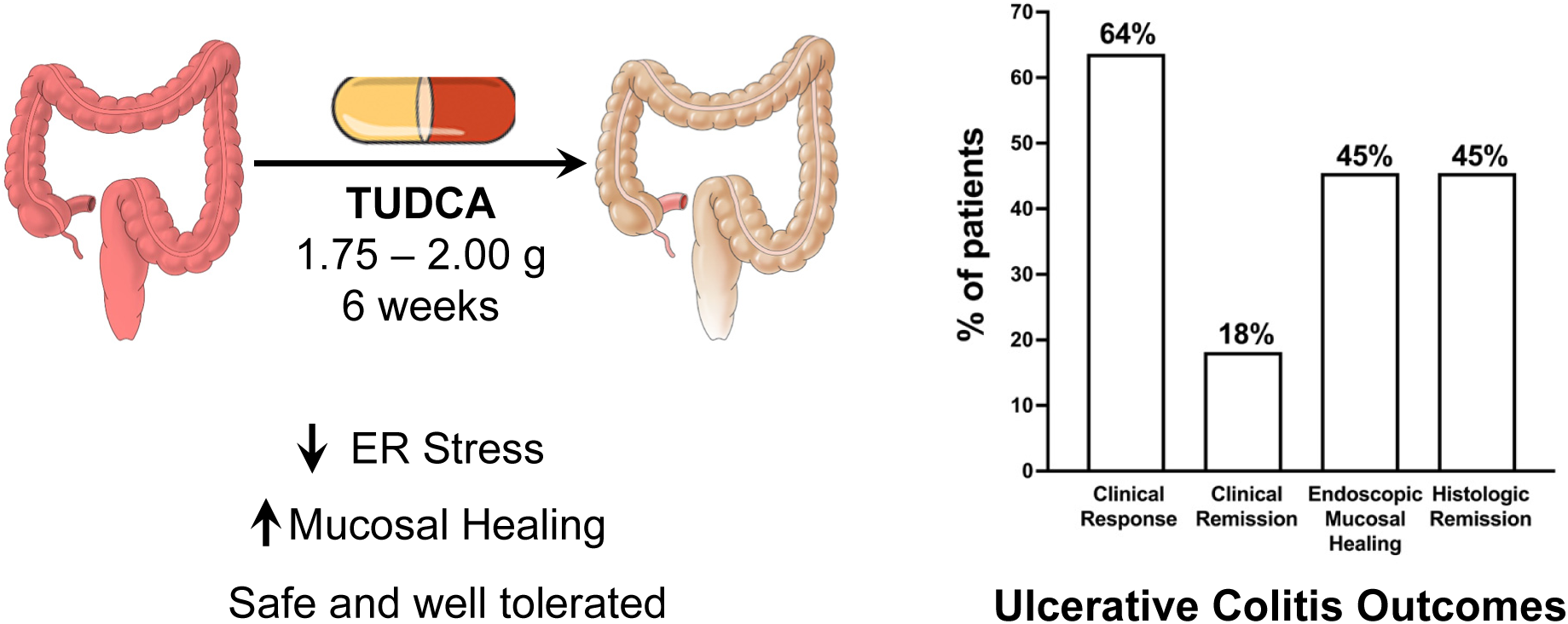

## INTRODUCTION

Ulcerative colitis (UC) is a form of chronic inflammatory bowel disease (IBD) that affects more than 1.2 million people in the United States and millions more worldwide.^1, 2^ Symptoms can be debilitating and include frequent loose stools, bloody diarrhea, and fecal urgency. Modern medical therapies include biologic drugs and small molecules inhibitors that are directed at reducing inflammatory signaling within the colon. Despite recent advances in these therapies, many patients experience persistently active colitis and 10-15% of patients have refractory disease or complications that necessitate a colectomy.^3–6^ Moreover, most of these therapies also cause systemic immune suppression and its inherent risks. Thus, there remains an unmet need for new UC therapies.

The potential to identify new IBD therapeutics rests on advancement in understanding of IBD pathophysiology. Decades of IBD research implicated uncontrolled mucosal immune responses and this led to several new and effective therapies that target inflammatory cytokines (TNFα, IL-23), immune signaling (Jak kinase inhibitors, immunomodulators), and immune cell trafficking (anti-integrins and S1P receptor modulators).^6^ More recent studies of genetics, the microbiome, and other environmental triggers have highlighted the critically important role of the intestinal epithelium in maintaining mucosal homeostasis and preventing IBD pathophysiology.^7^ To date, however, there are no IBD therapies that directly target the maintenance and repair of epithelial barrier function.

Unresolved protein misfolding in the epithelial endoplasmic reticulum (ER stress) and activation of the unfolded protein response (UPR) has emerged as an exciting potential therapeutic target in IBD. These cellular responses are activated in the inflamed ileal and/or colonic epithelium and play fundamental roles in epithelial barrier dysfunction and disease development.^8–15^ Human genetic studies also link disease risk with several genes associated with ER protein folding including *XBP1*, *AGR2*, and *ORMDL3*.^9, 16–19^ In IBD, oxidative stress, nutrient deprivation, increased protein synthesis rates, pathogenic microbiota, and inflammatory stimuli all contribute to disrupt ER protein folding homeostasis. These cause accumulation of unfolded or misfolded proteins and the resulting ER stress activates the UPR.^9, 12, 20–22^ The UPR is an adaptive response that eliminates misfolded protein in the ER through three mechanisms. First, the UPR transiently attenuates protein synthesis by PERK-mediated phosphorylation of the alpha subunit of eukaryotic translation initiation factor 2 (eIF2α). Second, activation of the UPR sensor IRE1 initiates splicing of XBP1 mRNA to produce an active transcription factor. Finally, The UPR sensor ATF6 traffics to the Golgi for cleavage by site-1 and site-2 processing enzymes that yield a soluble p50 fragment that migrates to the nucleus to activate transcription. The UPR induced genes encode molecular chaperones to assist protein folding, ER-associated degradation (ERAD) to eliminate misfolded protein, and trafficking machinery to assist protein cargo traffic out of the ER to the Golgi for secretion. If these adaptive mechanisms cannot resolve the protein-folding defect, cells enter apoptosis and epithelial barrier function is impaired.^23^

Our group found that ER protein misfolding, caused by deletion of genes which promote ER protein folding or molecular chaperone genes, activates the UPR (P58^IPK^/Dnajc3, ATF6) and increases sensitivity to experimental colitis in preclinical models.^10^ We further found that oral delivery of the chemical chaperone tauroursodeoxycholic acid (TUDCA) dramatically decreased the clinical, histological and biochemical signs of inflammation in both innate immunity- and T cell-dependent colitis through reducing ER stress in colonic intestinal epithelial cells (IECs). In other studies, TUDCA was also shown to antagonize TNFα mediated downregulation of nuclear receptors, improve bile acid homeostasis, combat dysbiosis, and abrogate experimental colitis and ileitis.^24–26^

TUDCA is a naturally occurring, hydrophilic secondary bile acid that has been used for centuries in Chinese medicine. The safety profile of TUDCA was demonstrated across several patient populations (obesity, diabetes, neurodegenerative disease) and it has been suggested as a possible novel therapeutic approach to IBD.^27–33^ In this study, we sought to translate the preclinical findings in colitis to humans by conducting a translational, open label trial with oral TUDCA supplementation with moderate to severely active UC. The primary endpoint was to determine the change of ER stress markers in endoscopically obtained biopsies before and after six weeks of treatment. We further evaluated safety, tolerability, and efficacy. Our findings demonstrate that TUDCA is well-tolerated, significantly reduces UC disease activity, effectively promotes mucosal healing, ameliorates inflammation, and reduces ER stress.

## RESULTS

### Patients

The study protocol including timing for endoscopy, surveys, disease activity assessment, and biospecimen acquisition is outline in Figure S1. Fourteen participants were screened and all enrolled in the study (Figure S2). The study population was mostly white (77%) with a female predominance (69%). Most patients had disease above the rectum (77%). At enrollment, all patients had moderate to severely active UC (total Mayo score of ≥6), with a median Mayo score of 9 and fecal calprotectin of 382 at baseline (Table 1 and 2). One patient withdrew consent at day 5 to pursue colectomy. Thirteen participants completed the study and were assessed for safety and tolerability. Two of the thirteen patients had study protocol deviations (discontinued mesalamine) leaving 11 participants assessable for clinical efficacy per protocol. All patients were on at least one concurrent UC medication including 9 (69%) who were on advanced therapy beyond mesalamine (Figure S3 and Table 2).

**Table 1.** Demographic and Baseline Clinical Characteristics of Study Participants.

|  |  |
| --- | --- |
| Characteristics |  |
| **Age (years), [range]** | 40.3 [20-63] (n=13) |
| **Sex, n (%)** |  |
| Female | 9 (69.2%) |
| Male | 4 (30.8%) |
| **Racial/ethnic Group, n (%)** |  |
| White | 10 (76.9%) |
| Black | 2 (15.4%) |
| Asian | 1 (7.7%) |
| **Disease Duration (years), [range]** | 13.2 [1.5-28] (n=13) |
| **Location (Montreal classification), n (%)** |  |
| E1 Proctitis | 3 (23.1%) |
| E2 Left Sided | 5 (38.5%) |
| E3 Extensive (pancolitis) | 5 (38.5%) |
| **Concurrent Meds (per patient), n (%)** |  |
| Prednisone | 2 (15.4%) |
| Infliximab | 2 (15.4%) |
| Ustekinumab | 2 (15.4%) |
| Vedolizumab | 5 (38.5%) |
| Mesalamine | 10 (76.9%) |
| Tofacitinib | 2 (15.4%) |
| Azathioprine | 2 (15.4%) |
| **Total # of UC Med Classes at Enrollment, n (%)** |  |
| 1 | 5 (38.5%) |
| 2 | 5 (38.5%) |
| 3 | 3 (23.1%) |
| **Total UCEIS (Mean, Median, Range)** | 5.0, 5, [1-8] |
| **Total Mayo score (Mean, Median, Range)** | 8.9, 9, [6-12] |
| PGA | 1.85, 2, [1-3] |
| Endoscopic | 2.38, 3, [1-3] |
| Rectal Bleeding | 1.92, 2, [0-3] |
| Stool Frequency | 2.69, 3, [1-3] |
| **Robarts Histopathology Index (Mean, Median, Range)** | 6.6, 6, [1-11] |
| **Fecal Calprotectin (Mean, Median, Range)** | 494.5, 382, [15.6-1000] |

**Table 2.**
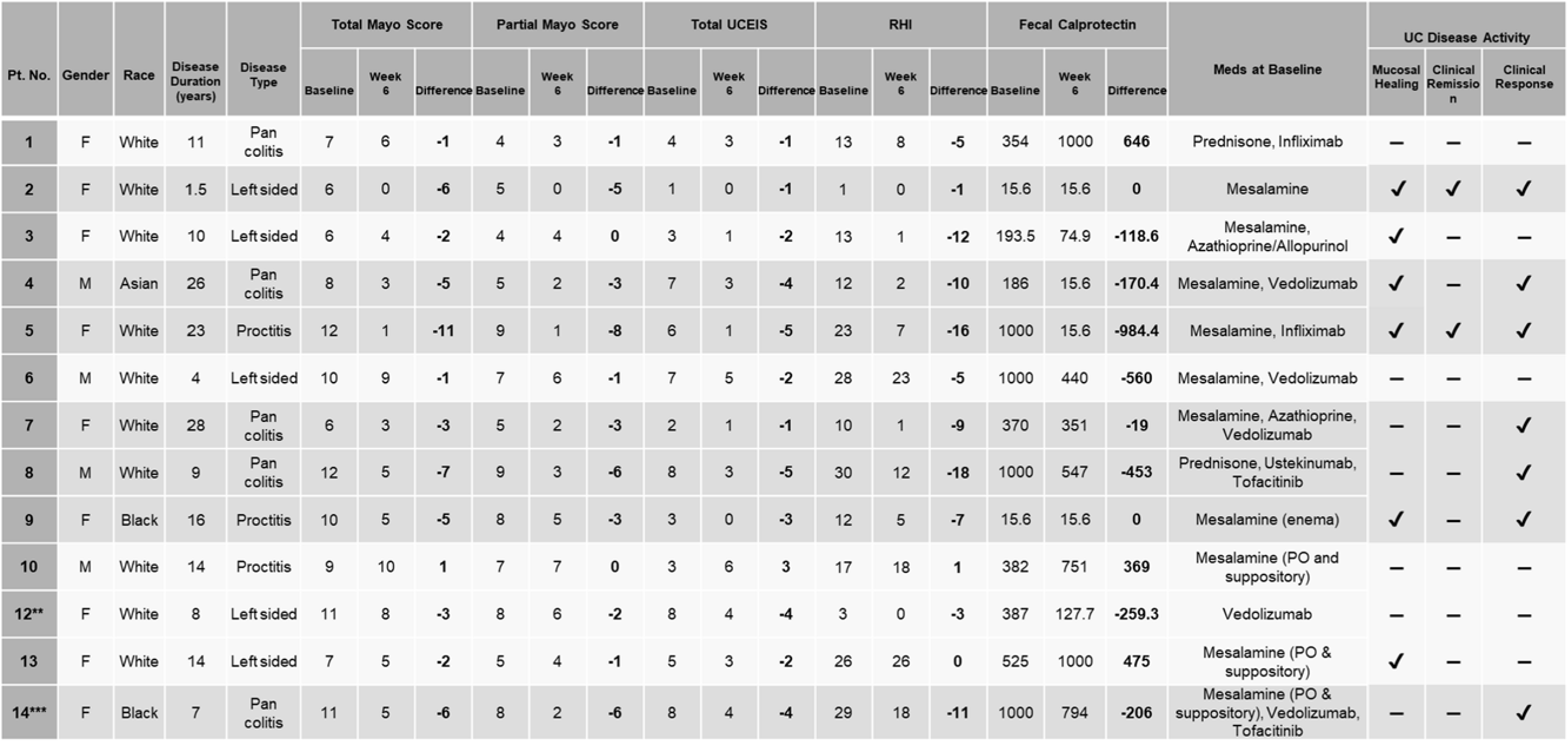
Baseline and Week 6 Data of Individual Study Participants. Patients highlighted in darker gray are clinical responders. Patients 10 and 13 stopped rectal mesalamine within two weeks of starting the study, which was a protocol deviation. * Patient #11 elected to come off protocol within one week to pursue a total abdominal colectomy. ** Patient #12 Dose reduced to 1000 mg/day for 8 days, then returned to 1750 mg/day. *** Patient #14 Dose reduced to 1000 mg/day for 69 days.

### Safety and Tolerability

In this UC trial cohort, TUDCA therapy was safe and well-tolerated with no serious adverse events (SAE; those which would pose a threat to patient life or ability to function). All adverse events (AE) potentially related to TUDCA were gastrointestinal in nature, transient, and classified as mild or moderate with a CTCAE Grade 1 or 2 (Table S1). Of the 13 patients who took TUDCA for the study duration, nausea or dyspepsia was the most common AE occurring in 6 patients (46%). All cases had symptomatic resolution within 5 days. A mild increase in diarrhea (1-2 extra bowel movements per day) occurred in two patients lasting for 3 and 14 days. Two patients reduced their TUDCA dose from 1750 mg to 1000 mg for nausea or diarrhea. Patient 12 resumed their starting dose (1750 mg) after 1 week and tolerated this well. Patient 14 tolerated 1000 mg well and stayed on this for the duration of the study. TUDCA administration was not associated with any significant changes in kidney or liver function tests, nor in cell blood counts (Table S2).

### Clinical Outcomes

We used the Mayo score to assess UC activity. Patients taking TUDCA per trial protocol exhibited significant improvements in clinical and endoscopic disease activity at week 6. From baseline to week 6, these patients exhibited significant improvements in all component measures of the total Mayo score including stool frequency (p=0.03), rectal bleeding (p=0.002), endoscopy (p=0.02), and physician’s global assessment (p=0.03) (Figures 1A-D), as well as total Mayo score (mean change 9 to 4.5; p=0.001) (Figure 1E). The partial Mayo score decreased at all time points (Figure 1F). In sum, six weeks of TUDCA induced clinical response, clinical remission, and endoscopic mucosal healing in 64%, 18%, and 45% (Figure 1G; definitions in figure legend). All patients also experienced improvements in the Ulcerative Colitis Endoscopic Index of Severity (UCEIS) and the Simple Clinical Colitis Activity Index (SCCAI) (Figure S4 and S5 A, B).

**Figure 1.**
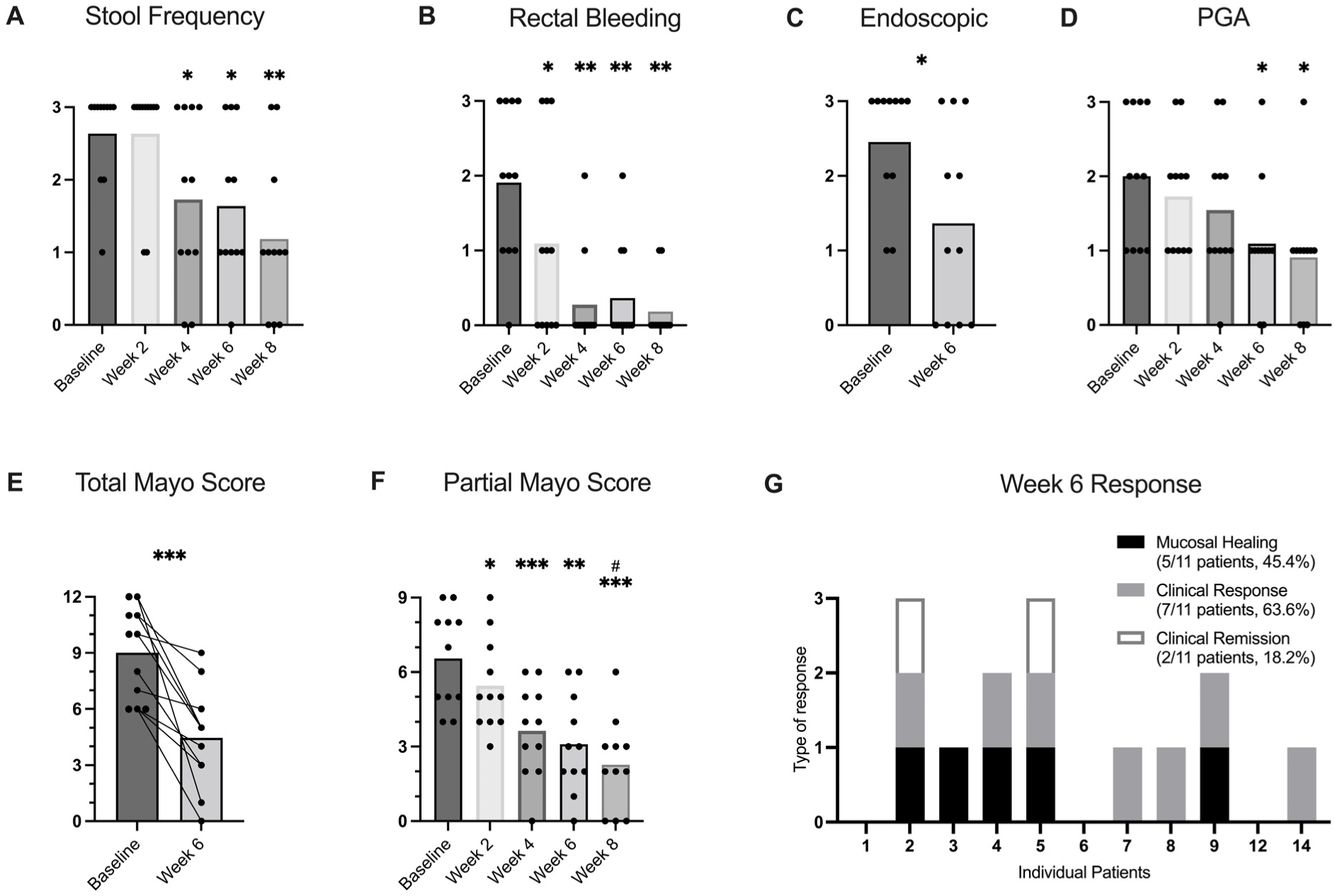
TUDCA reduces disease activity in patients with moderate to severely active ulcerative colitis. Mayo ulcerative colitis scoring index is provided for patients who completed TUDCA therapy per protocol (N=11). **Top**: Mayo subscore components. P-values *<.05, **<.01, ***<.001.P<0.001 by two-way ANOVA for all multiple time point measures (stool frequency, bleeding, PGA). **Bottom**: Total Mayo score, partial Mayo score (without endoscopic score), and individual patient responses at week 6 based on total Mayo score. P-values *<.05, **<.01, ***<.001. # P = 0.03 comparing week 6 and week 8. P<0.001 by two-way ANOVA for Partial Mayo Score. The following definitions were used. Endoscopic Mucosal Healing: a decrease of at least 1 point on the Mayo endoscopic subscore and final score of 0 or 1. Clinical Response: A decrease in Mayo Score of ≥ 3 points and a decrease in Mayo Score of ≥ 30% from baseline and a decrease in the rectal bleeding score of ≥ 1 or an absolute rectal bleeding score of 0-1. Clinical Remission: Total Mayo Score of ≤2 with no subscore >1 and improvement of endoscopic appearance of the mucosa (Mayo 0 or 1).

The effect of TUDCA was both rapid and sustained in the short term after withdrawal. Within two weeks of starting TUDCA, patients demonstrated significant decreases in rectal bleeding and partial Mayo score (Figures 1B and 1F). Stool frequency was significantly reduced by four weeks (Figure 1A). Partial Mayo scores (without endoscopic subscore) decreased further two weeks after TUDCA withdrawal from week 6 to week 8 (Figure 1F, p=0.03). This improvement was driven by lower rectal bleeding and stool frequency subscores, suggesting ongoing benefit after TUDCA was stopped. Overall, patients experienced an increase in IBD related quality of life over the study course as evidenced by increases in SIBDQ scores in 10 of 11 patients (Figure S5 C). Though the cohort was small, no findings differed by sex. Compared to the SPARC-IBD institutional control cohort, patients on TUDCA were less likely to have started a new UC therapy or increased their current therapy at 6 months (27% vs 77%, p<0.01) (Table 3 and 4).

**Table 3.**
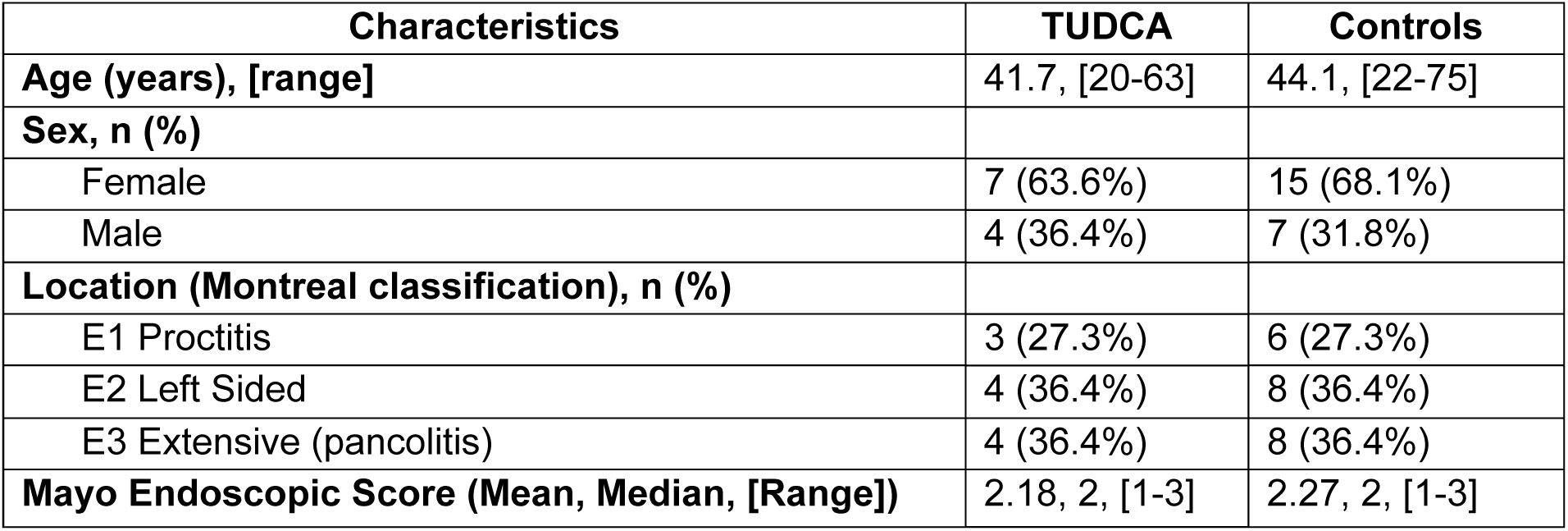
Demographics and Baseline Clinical Characteristics of TUDCA treated patients and matched institutional control cohort. Baseline characteristics of TUDCA clinical trial patients (n=11 evaluable per protocol) and retrospective controls derived from SPARC-IBD database (n=22, 2:1 matching).

**Table 4.**
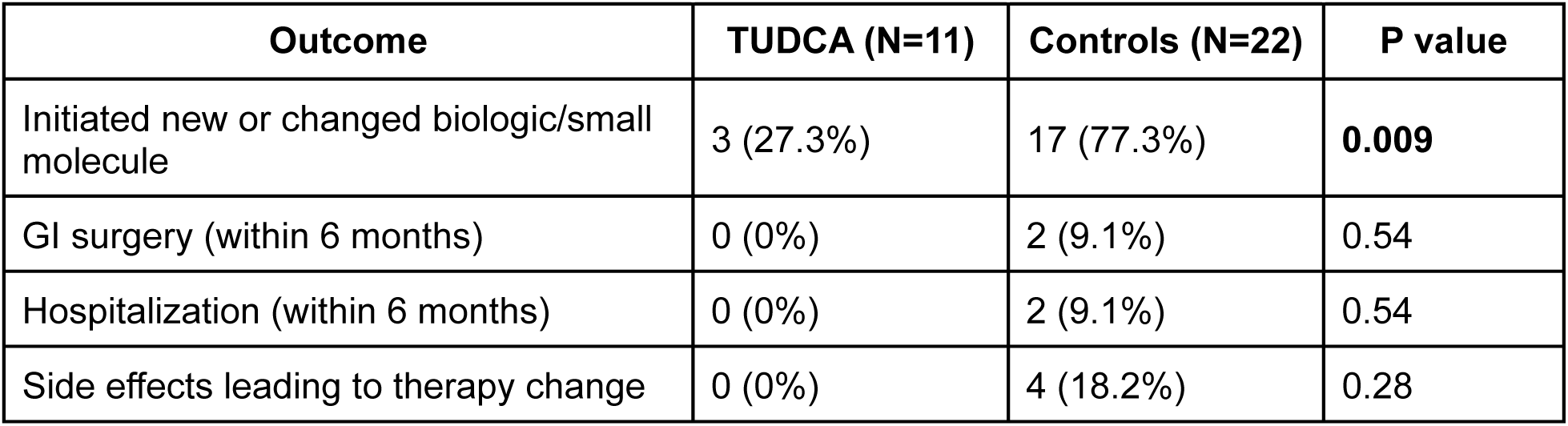
Clinical Outcomes at 6 months in patients treated with TUDCA versus matched institutional cohort. Comparison of clinical outcomes at 6 months between TUDCA patients and controls Statistics by Fisher’s exact test (two-sided).

| Outcome | TUDCA (N=11) | Controls (N=22) | P value |
| --- | --- | --- | --- |
| Initiated new or changed biologic/small molecule | 3 (27.3%) | 17 (77.3%) | **0.009** |
| GI surgery (within 6 months) | 0 (0%) | 2 (9.1%) | 0.54 |
| Hospitalization (within 6 months) | 0 (0%) | 2 (9.1%) | 0.54 |
| Side effects leading to therapy change | 0 (0%) | 4 (18.2%) | 0.28 |

### Markers of Mucosal Repair and Restitution

At week six, 45% of patients achieved histologic remission (RHI ≤ 2; Figure 2A). Fecal calprotectin (FCP) was compared from baseline to week 6 (Figure 2B and Table 2). Of the nine patients who had FCP levels greater than 150 mcg/g, all but one exhibited a decrease in FCP.

**Figure 2.**
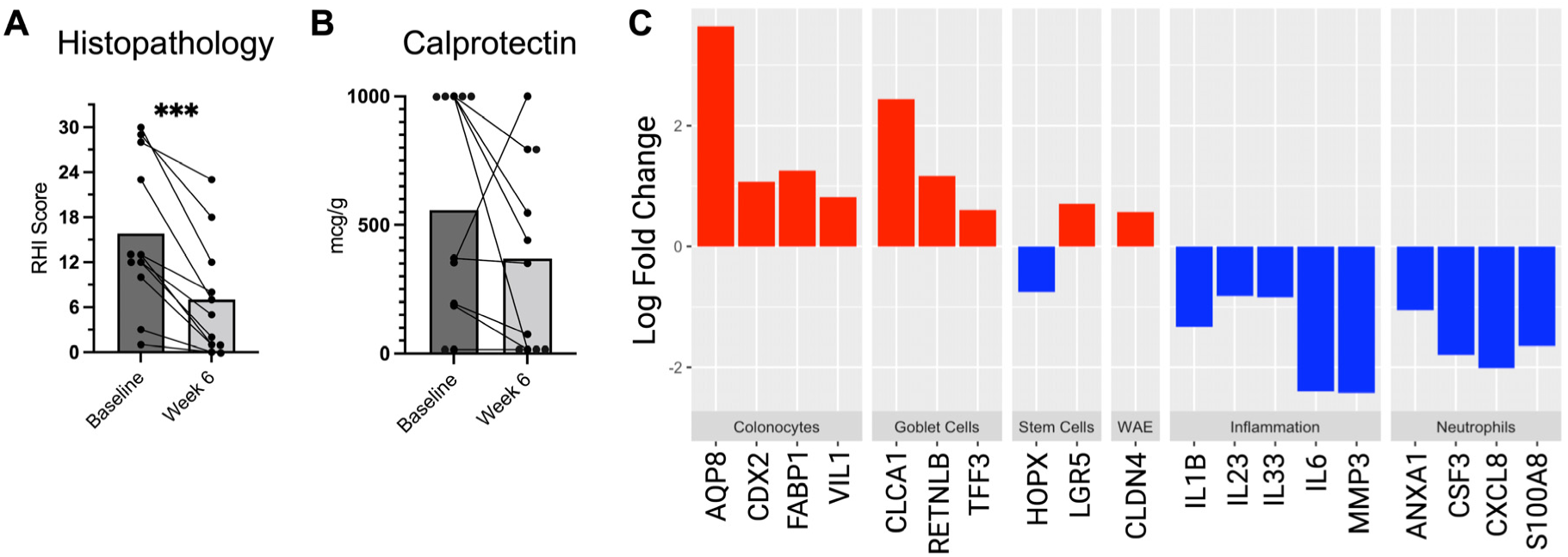
TUDCA promotes histologic healing and mucosal repair. **A)** Histologic disease activity using the Robart’s Histopathology Index (RHI). **B)** Fecal calprotectin. **C)** Change in mucosal expression of genetic markers related to inflammation and barrier disruption. Positive log fold change is represented in red and negative log fold change blue. All genes are significant with *P ≤ 0.05*.

Mucosal biopsy samples were subjected to gene expression profiling by RNA-Seq and examined for differences after 6 weeks of TUDCA treatment. Post treatment biopsies demonstrated a pattern consistent with mucosal healing with an increase in markers of mature colonocytes and goblet cells and a decrease in markers of inflammation and neutrophil infiltration (Figure 2C and S6). The expression data also revealed evidence of epithelial repair and regeneration as reduction in colitis-associated regenerative stem cells (*HOPX*) was coupled with an increase in homeostatic stem cells (*LGR5*) and the wound-associated epithelial cells (*CLDN4*) that form the initial barrier over intestinal ulcers (Figure 2C).^34–37^ Finally, there were significant decreases in cytokines associated with inflammation in IBD including IL-23, IL-6, and IL-1β.

Pathway analysis using the KEGG database also revealed a pattern consistent with resolving UC inflammation and mucosal healing (Figure 3) This included increased expression of pathways characteristic of having an intact epithelium, such as metabolic, absorptive, and fatty acid degradation pathways. There was also a reduction in pathways associated with UC activity including chemokine signaling, cytokine-cytokine interactions, T-cell receptor activation, NF-κB, as well as osteoclast differentiation.^38–43^ Notably, Th-17 cell differentiation, IL-17 signaling, and IgA production were all also decreased. These findings are consistent with recent reports identifying intestinal epithelial ER stress promotes Th17 cell differentiation and orchestrates IgA responses in the gut.^44, 45^

**Figure 3.**
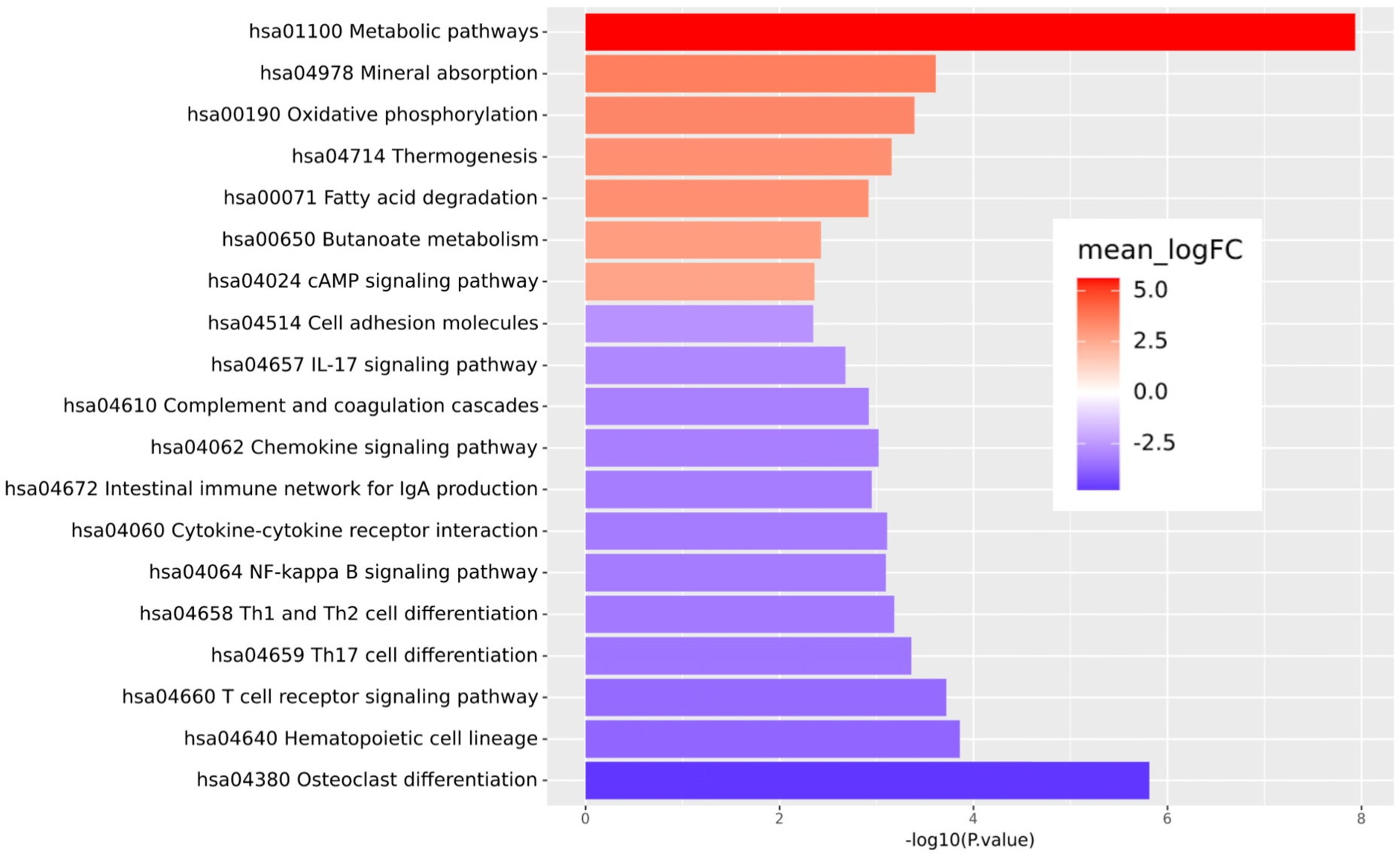
Pathway analysis reveals pattern of mucosal gene and reduced inflammation. Gene expression analysis of mucosal biopsies was performed by RNA-Seq on all pre- and post-TUDCA treatment mucosal biopsy specimens. Top upregulated (red) and downregulated (blue) pathways using the Kyoto Encyclopedia of Genes and Genomes (KEGG) database.

### Reduction in ER stress and the UPR

TUDCA treatment led to significant reductions in mucosal expression of multiple ER stress and UPR genes. A heatmap shows individual expression data that is further divided by clinical responders and non-responders (Figure 4A). Notable reductions were found in *DNAJB9* (*HSP40*), *ATF4*, and *BiP (HSPA5). DNAJB9*, was the most highly downregulated gene and is a transcriptional target for spliced XBP1, which links ER stress to intestinal inflammation and is a genetic risk factor for IBD.^9^ Finally, immunohistochemical staining for BiP revealed that expression levels positively correlated with histopathology scores and were reduced after 6 weeks of oral TUDCA (Figure 4 B,C,D).

**Figure 4.**
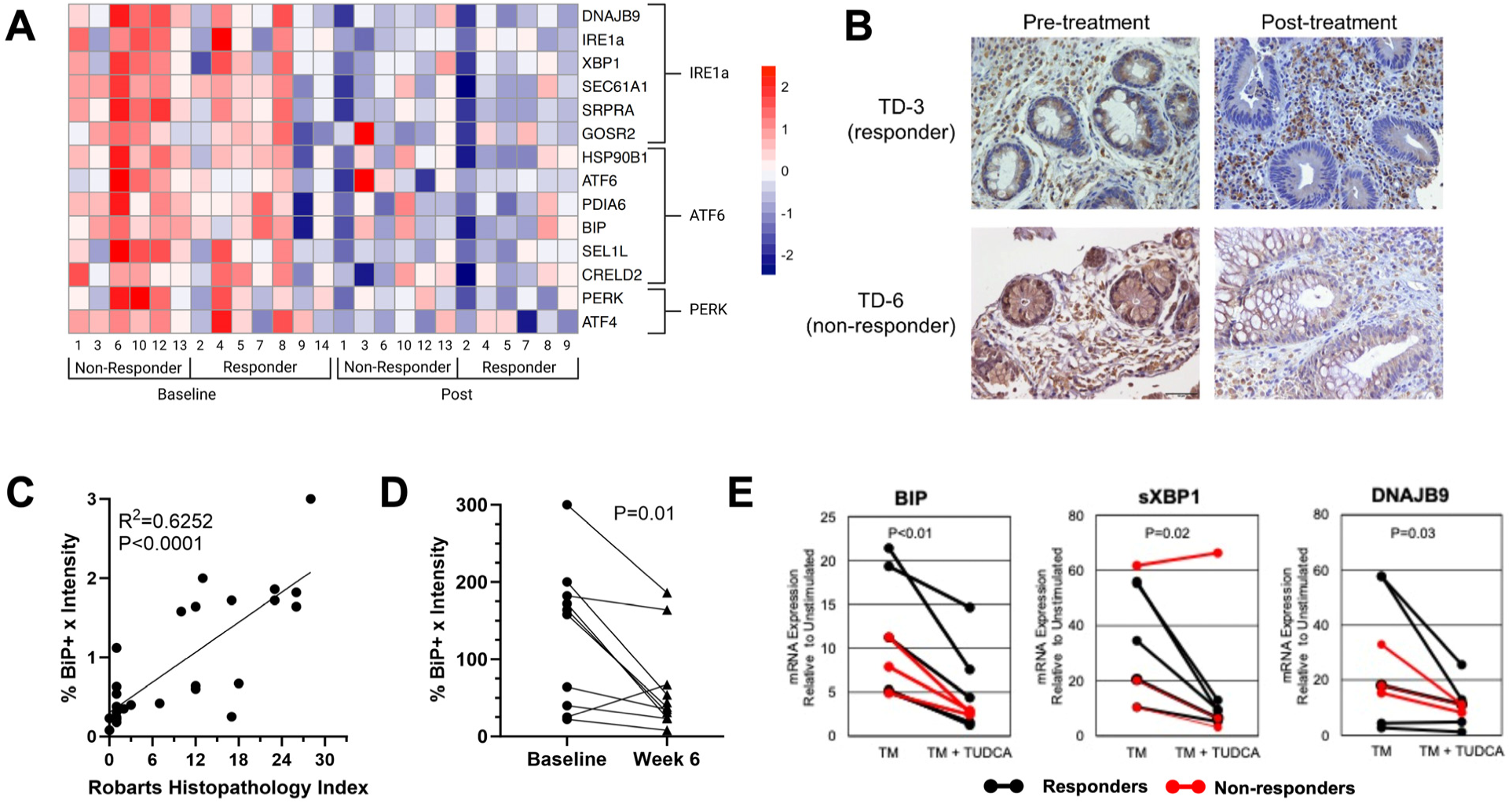
TUDCA reduces epithelial ER stress and the unfolded protein response *in vivo* and *ex vivo*. **A)** The RNA-seq data was queried for genes expression changes falling within ER stress and UPR pathways *IRE1a*, *ATF6*, and *PERK*. Data for individual patients are represented and separated by clinical responders and non-responders. All genes represented have significance of P<0.05. P=NS between the responder and non-responder cohorts. **B)** Representative immunohistochemistry for BiP on pre and post treatment biopsies. **C)** Positive correlations between the BiP immunohistochemistry score (percent positive cells x intensity on a 0-3 score) and histopathology. **D)** Reduction in the percent positive BiP epithelial cells before and after TUDCA treatment. Statistics performed by simple linear regression and Student’s t-test. **E)** Patient derived colonic organoids were differentiated for 72 and stimulated with vehicle control, tunicamycin (TM; 1 ug/ml), or TM after 1-hour pretreatment with TUDCA (5 mM). mRNA expression of ER stress response genes BIP, spliced-XBP1, and DNAJB9 were examined by RT-PCR. Data is graphed relative to vehicle control mRNA levels for individual cell lines.

Colonic organoids were also derived from mucosal biopsies to test for *ex vivo* response to TUDCA therapy. Three clinical non-responders and five clinical responders to TUDCA were each used to grow a respective organoid cell line. Successfully derived organoid lines were subjected to ER stress inducer tunicamycin (TM) in the presence or absence of TUDCA and evaluated for expression of UPR genes using qRT-PCR. Though the amplitude of change varied across cell lines, tunicamycin consistently increased expression of *BiP*, *sXBP1*, and *DNAJB9* while TUDCA treatment reduced evidence of ER stress (Figure 4E). Altogether, these results confirm across a population of patients with UC that TUDCA reduces colonic mucosal and epithelial ER stress.

### Microbiome and bile acid biomarkers of response

The intestinal microbiome and bile acids are highly connected to one another as well as to regulation of mucosal homeostasis in IBD. We performed 16S sequencing on stool and evaluated serum bile acids at baseline and after 6 weeks of TUDCA. While the total amount of serum bile acid concentration did not change with TUDCA treatment, there were shifts in distribution including increases in deoxycholic acid (DCA), ursodeoxycholic acid (UDCA) and lithocholic acid (LCA) and compensatory decreases in the proportions of taurocholic acid (TCA) and glycocholic acid (GCA) (Figure 5A). The change in serum TUDCA levels positively correlated with change in clinical disease activity as measured by the total Mayo score, suggesting this is potentially a biomarker of response (Figure 5B).

**Figure 5.**
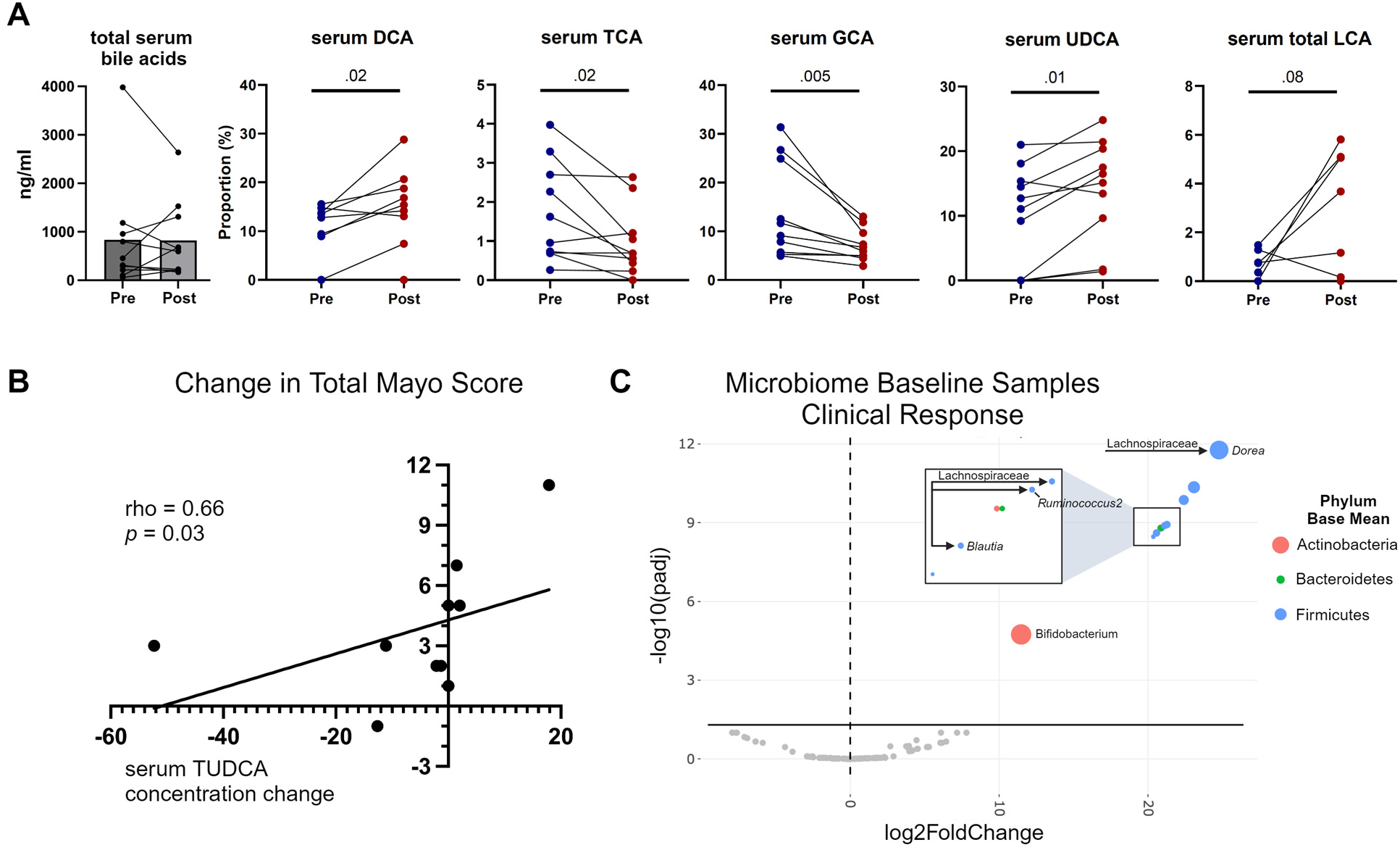
Changes in serum bile acid and microbiome composition with TUDCA treatment. **A)** Serum bile acids (LC-MS/MS) analysis revealed shifts in distribution following TUDCA treatment with increases in DCA, UDCA and LCA and compensatory decreases in the proportions of TCA and GCA. **B)** Changes in serum TUDCA (ng/ml) correlate with improvements in total Mayo score. **C)** The baseline microbial composition in patients with clinical response and/or mucosal healing is enriched in specific bacterial taxa verses non-responders. All circles in color and above solid horizontal black line represent those with significant (p<0.05) differential expression.

The stool microbiome was examined for changes with therapy and for the identification of potential biomarkers of response. Overall alpha and beta diversity of the microbiome did not change from pre to post therapy; however, composition shifts were apparent when divided by responders and non-responders (Figure S7). Biomarker analysis revealed that pretreatment stool from patients achieving clinical or endoscopic response was enriched in *bifidobacterium* and *lachnospiraceae* as compared to non-responders (Figure 5C). Several of these *lachnospiraceae* (*dorea*, *blautia*, and *ruminococcus*) are linked to mucosal homeostasis and UC remission.^46–48^ After treatment, further increases in *bifidobacteria*, *clostridium* species IV, and *ruminococcus* were observed in responders as compared to non-responders (Figure S8A). Finally, while responders showed specific microbial changes pre-versus post-treatment, non-responders did not (Figure S8B,C). These findings suggest that microbiome may shape the response to TUDCA and that the presence of certain bacterial species may serve as positive predictors of clinical effect.

## DISCUSSION

Patients with moderate-to-severely active UC who are refractory to modern biologic or small molecule therapy have limited treatment options beyond surgery. In this open label, translational clinical trial, we tested the approach of adding an orally available drug, tauroursodeoxycholic acid (TUDCA), to a patients’ existing therapeutic regimen. TUDCA was rationally selected based on its capacity to mitigate unresolved ER stress in the intestinal epithelium, a recognized contributor to IBD pathophysiology. As a primary endpoint, we found that 6 weeks of TUDCA led to a reduction in markers of ER stress and inflammation in mucosal biopsies. TUDCA was safe and well-tolerated. Clinically, patients taking TUDCA experienced objective improvements in clinical, endoscopic, and histologic disease activity.

There is a well-recognized ceiling of efficacy for current immunosuppressive IBD therapies.^49, 50^ For biologics, initial 8-week response rates are reasonable (53-67%), but 1-year net remission rates are low (22-35%). In patients previously exposed to another biologic, the outcomes are worse for both response (37-52%) and remission (10-21%). Oral JAK inhibitors are slightly better in biologic exposed patients, but still only achieve long term remission rates of 10-33% and typically require that patients stay on the highest drug dose. Considering the 8 patients in this trial who were already taking at least one biologic, TUDCA therapy led to 6-week clinical response of 63%. Moreover, 77% of patients who took TUDCA for only 6 weeks did not require a further change or escalation of therapy at 6 months as compared to 23% of an institutional control cohort. This suggests that a therapy targeting epithelial barrier dysfunction may help break the therapeutic ceiling in UC.

ER stress is documented to play a role in the etiology of diabetes and a clinical trial demonstrated that TUDCA increased insulin sensitivity by 30% (p<0.05).^51, 52^ In contrast, here we show TUDCA treatment for UC reduces UC symptoms, inflammation, and ER stress markers by ∼50% (<0.001). The greater efficacy may result from greater TUDCA access to the intestine by oral delivery, the nature of the etiology of the disease or the significance of protein misfolding in the ER as a main factor in the disease pathogenesis. Further large-scale placebo-controlled studies are required to test these notions.

There are multiple mechanisms by which TUDCA treatment may promote mucosal healing. A decrease in ER stress and UPR activation was observed in the epithelium and barrier repair was evidenced by histologic improvement, increases in mature colonocyte markers, and restitution of metabolic pathways. Objective reductions were observed in markers of innate and adaptive immune activation including cytokine signaling (*e.g.,* IL6, IL17, IL23, NFĸB), indicating that TUDCA promoted immunologic mucosal homeostasis as well. The possibility that TUDCA directly shapes the immune cell compartment in UC is supported by evidence that the UPR influences T cell differentiation, activation, and function.^53^ However, some immune effects may relate back to epithelial ER stress which is known to promote Th17 cell differentiation and orchestrate IgA responses.^44, 45^ Finally, increased mucus secretion by goblet cells may be another mechanism by which TUDCA promotes barrier function and homeostasis as alleviation of ER stress promotes a thicker mucus layer, shapes mucosal associated microbiota, and protects from experimental colitis in a NOD2 dependent manner.^54^

It is important to consider how TUDCA might be added to our current armamentarium of UC therapies. A strength of this study was the intentional inclusion of patients taking a variety of medication regimens and with varied disease extent (proctitis to pan-colitis). Given its strong safety profile across a variety of disease states, unique mechanism of action, and relatively low price point as compared to biologics, a practical approach may be to combine TUDCA with immunosuppressive biologic and small molecule UC therapies. Indeed, the trial data showed that TUDCA reduced ER stress and UC clinical symptoms in patients regardless of concurrent therapy. As incomplete response is common with current therapies, a future placebo-controlled trial might be designed to study it as a new addition to patients flaring on a current therapy. Alternatively, TUDCA may be able to boost remission rates when started concurrently with TNFα inhibitor induction therapy for example. This combination approach is currently under therapeutic evaluation in Wolfram syndrome (NCT05676034), a rare genetic disorder linked to ER dysfuncton and stress.^55^ Finally, the choice of TUDCA may be guided by microbial and/or serum biomarkers if the observations herein held true in a larger study population.

Limitations of the current trial may be used to inform the design of a phase II trial. This trial was a single arm, open label trial with a primary endpoint of change in ER stress at 6 weeks. The subsequent trial should be placebo controlled with clinical and endoscopic improvement as primary endpoints. Consideration should also be made to extend the treatment course to 8 or 12 weeks given the finding that partial Mayo scores continued to improve even after TUDCA was stopped. Eventually, larger trials could be designed to study continued TUDCA therapy as a maintenance medication once remission is achieved. Further exploration for biomarkers that predict treatment success should also be part of a larger trial and may include *ex-vivo* approaches (organoids), pre-treatment microbiome, or biopsy assessments for ER stress markers such as BiP.

In this six-week open label trial, oral TUDCA was well tolerated, met the primary endpoint of reducing ER stress, and improved UC activity in patients who were unresponsive to their current therapy. Prior studies support the existence of a mechanistic link proposed between protein misfolding in the ER of IECs and development of IBD in humans^10^. The simplicity of oral TUDCA and the novel approach to restoring the epithelial barrier in UC underscores the important need for a randomized controlled trial evaluating TUDCA.

## METHODS

### Study Overview

This was a translational, open-label and non-randomized, clinical trial conducted at one academic medical site (Washington University School of Medicine, St. Louis, MO) from 2019 to 2022. Trial registration can be found at clinicaltrials.gov (NCT04114292). This study is reported according to the Transparent Reporting of Evaluations with Nonrandomized Designs (TREND) statement.^56^

In this investigator initiated clinical trial, pharmaceutical grade tauroursoursodeoxycholic acid (Tudcabil^TM^) was obtained from Bruschettini Srl, (Genoa, Italy). Capsules with 250 mg TUDCA were provided in foil packaging and contained other constituents limited to microcystalline cellulose, lactose, corn starch and magnesium stearate. The study was reviewed and approved by the FDA under an Investigational New Drug Application (IND 142064).

### Sex as a Biological Variable

Our study examined male and female patients, and similar findings are reported for both sexes.

### Patients and Enrollment Criteria

Key inclusion criteria included patients who were 18 to 65 years of age with evidence of moderate to severe UC (Mayo score ≥6, endoscopic subscore ≥1). Patients were on a stable dose of IBD medications including no change in biologic, small molecule or mesalamine therapy within 8 weeks of study enrollment and a stable or decreasing dose of corticosteroids for at least 4 weeks. As TUDCA addresses a different mechanisms of action, this study intentionally included a heterogeneous UC population with regard to disease extent and concurrent therapy. Key exclusion criteria included chemical chaperone therapies in the 3 months prior to screening, cholestatic pathologies, bile acid binding therapies, pregnancy/lactation, and baseline liver transaminases (AST or ALT) > 1.5 times the upper limit of normal. Full inclusion and exclusion criteria can be found on NCT04114292.

### Study Design and Outcome Measures

The overall experimental design is summarized in Supplementary Figure 1. Participants received oral TUDCA (1750-2000 mg/day dosed TID with 250 mg capsules) as an adjunctive intervention to standard medical care. As TUDCA is not immunosuppressive, all concurrent IBD therapies were allowed. A colonoscopy or flexible sigmoidoscopy with biopsy, blood draws, stool collection, questionnaires, and a clinical exam were completed at baseline (typically the day before starting TUDCA) and after 6 weeks of therapy on their last day of therapy. Phone based follow up was performed at weeks 2, 4, and 8 (two weeks after stopping TUDCA). Chart review was completed for each patient at months 2, 4, 6, and 12 to monitor clinical course. The Common Terminology Criteria for Adverse Events (CTCAE) v5.0 was used to assess safety and tolerability.

The primary endpoint was a significant decrease in ER stress markers in mucosal biopsies. Secondary end points included monitoring safety and tolerability, change in total Mayo score, and histologic improvement using the Robarts Histopathology Score (range 0-33).^57^ The Mayo score for UC disease activity (range 0-12) is the sum of four component subscores (range 0-3) including diarrhea, rectal bleeding, endoscopic severity, and physicians global assessment.^58^ The partial Mayo score includes all components except for endoscopy.^59^ The following definitions were used for outcomes: Endoscopic Mucosal Healing: a decrease of at least 1 point on the Mayo endoscopic subscore and final score of 0 or 1. Clinical Response: A decrease in Mayo Score of ≥ 3 points and a decrease in Mayo Score of ≥ 30% from baseline and a decrease in the rectal bleeding score of ≥ 1 or an absolute rectal bleeding score of 0-1. Clinical Remission: Total Mayo Score of ≤2 with no subscore >1 and improvement of endoscopic appearance of the mucosa (Mayo 0 or 1).

As this was an open label, single-arm trial, we queried the Crohn’s and Colitis Foundation’s SPARC-IBD database of patients enrolled at Washington University to develop an institutional “control cohort” of patients that were matched (2:1 SPARC:TUDCA trial) for age, sex, extent of disease and Mayo endoscopic subscore (Table 3 and 4).^60^ This control cohort was used to compare clinical outcomes at 6 months.

### Laboratory and Biologic Assessments

Routine hematology, chemistry, and fecal calprotectin (FCP) labs were performed at Barnes Jewish Hospital Laboratories. Biopsies were immediately separated into RNA later (Thermo Fisher), flash frozen, or cultured for organoids as previously described by our group.^61^ Robarts histopathological index (RHI) scoring was performed by an expert GI pathologist (author TL) blinded to patient identifiers and timepoints. Biologic comparisons were made between pre-and post-treatment biopsy samples from the same location of 10-15 cm from anal verge. RNA was isolated by RNAeasy kit (Qiagen) and sequencing (RNA-seq) performed by the Genome Technology Access Center at Washington University using Illumina NovaSeq 2×150 with a 30M read target. Bioinformatics included the R/Bioconductor package Limma using the voomWithQualityWeights function for differential expression analysis and the Kyoto Encyclopedia of Genes and Genomes (KEGG) database for pathway analysis.

Stool samples were processed for 16S rDNA Illumina sequencing and analysis. Primer selection and PCRs were performed as described previously.^62, 63^ DNA Sequencing was performed at the Innovation Lab (Washington University School of Medicine) for sequencing by the 2 × 250bp protocol with the Illumina MiSeq platform. Read quality control and the resolution of amplicon sequence variants were performed in R version 4.0.5 with DADA2 version 1.18.0 using the Ribosomal Database Project (RDP) trainset 16 release 11.5.^64, 65^ Ecological analyses were performed using PhyloSeq version 1.46.0 and additional R packages (version 4.3.2).^66^ Differentially abundant taxa were identified using DESeqs.^67^ 16S sequencing data have been uploaded to the European Nucleotide Archive.

### Statistical Considerations and Analyses

Sample size calculations were determined based on results from preclinical modeling where ER stress marker RNAs including sXBP1 and BIP were significant reduced by 25% to 45% after TUDCA treatment. .^10, 68^ Using alpha set to 0.05 and beta to 0.10 (90% power), power calculation showed that seven patients were sufficient to detect the difference in before and after treatment. We expanded the cohort to 14 to further assess tolerability as well as to account for variability in IBD patient disease activity. Specific statistics including 2-way ANOVA, Student’s paired t-test, non-parametric *Spearman,* and Wilcoxon signed-rank test were utilized as appropriate and are described in the figure legends. For all outcomes a two-sided p <0.05 was considered statistically significant. Statistical analyses were performed using SPSS, Prism, and R software. All authors had access to the study data, and reviewed and approved the final manuscript.

### Study Approval

The protocol was approved by the institutional review board (201812101) and all patients provided written informed consent.

## Supporting information

Supplementary Methods and Figures Legends JCI

## Data Availability

All data produced in the present work are contained in the manuscript

## Data Availability

Data, analytic methods, and study materials will be made available to other researchers. RNA-Sequence data will be made available on the Gene Expression Omnibus (GEO) database repository (GSE266963). 16S sequencing data has been uploaded to the European Nucleotide Archive (accession number PRJEB73763).

## Author Contributions

**Conceptualization:** VL, MW, NOD, RJK, MAC. **Methodology:** VL, KH, RD, SC, DD, NOD, RJK, MAC. **Software:** VL, MAC. **Validation:** VL, KH, RD, NOD, RJK, DD, MAC **Formal Analysis:** VL, KH, RD, SS, RR, NOD, RJK, MAC **Investigation:** VL, KH, RD, MW, TP, YX, TL, DD, GOE, SS, RR, NOD, RJK, MAC **Resources:** RJK, MAC **Data Curation:** VL, KH, DD, SS, RR, NOD, RJK, MAC **Writing – Original Draft:** VL, RJK, MAC **Writing – Review and Editing:** VL, RJK, RD, NOD, MAC **Visualization:** VL, RJK, MAC **Supervision:** MTB, NOD, RJK, MAC **Project administration:** RJK, MAC **Funding acquisition:** RJK, NOD, MAC.

## Acknowledgements

We appreciate critical review of the manuscript by Professor Fumihiko Urano MD, PhD and Dr. Gabriele Nicolini PharmD, PhD.

