## Supplementary Methods and Figures Legends JCI for "Tauroursodeoxycholic Acid (TUDCA) Reduces ER Stress and Lessens Disease Activity in Ulcerative Colitis"

### SUPPLEMENTARY METHODS AND MATERIALS

#### Immunohistochemistry

Formalin-fixed paraffin-embedded tissue blocks used for this study were collected at Washington University School of Medicine under an Institutional Review Board-approved study. Four-micron thick tissue sections were used for immunohistochemical staining. Sections underwent heat mediated antigen retrieval with citrate buffer at pH 6, then incubated with anti-human XBP1 (1:500) and anti-human HSPA5/BiP (1:100). The reaction was revealed using Novolink Polymer (Leica Microsystems) and the slides were counterstained with Mayer's Hematoxylin.

| Primer | Vendor | 5' -> 3' |
|---|---|---|
| UGT1A9 Forward | Integrated DNA Technologies | CAG GTT TTG TGC TGG TAT TTC TCC CA |
| UGT1A9 Reverse | Integrated DNA Technologies | GCG GAT ATC CAT AGG CAC TGG CTT TCC CTG ATG ACA |
| tXBP1 Forward | Integrated DNA Technologies | TGG CCG GGT CTG CTG AGT CCG |
| tXBP1 Reverse | Integrated DNA Technologies | ATC CAT GGG GAG ATG TTC TGG |
| sXBP1 Forward | Integrated DNA Technologies | CTG AGT CCG AAT CAG GTG CAG |
| sXBP1 Reverse | Integrated DNA Technologies | ATC CAT GGG GAG ATG TTC TGG |
| CHOP Forward | Integrated DNA Technologies | AGA ACC AGG AAA CGG AAA CAG |
| CHOP Reverse | Integrated DNA Technologies | TCT CCT TCA TGC GCT GCT TT |
| BIP Forward | Integrated DNA Technologies | TGT TCA ACC AAT TAT CAG CAA ACT C |
| BIP Reverse | Integrated DNA Technologies | TTC TGC TGT ATC CTC TTC ACC AGT |
| GRP94 Forward | Integrated DNA Technologies | GAA ACG GAT GCC TGG TGG |
| GRP94 Reverse | Integrated DNA Technologies | GCC CCT TCT TCC TGG GTC |
| GAPDH Forward | Integrated DNA Technologies | GTC TAG AAA AAC CTG CCA AAT ATG |
| GAPDH Reverse | Integrated DNA Technologies | CTG TTG AAG TCA GAG GAG ACC AC |

| Reagent Type | Name | Vendor | Catalog Number |
|---|---|---|---|

| | | | |
|---|---|---|---|
| Coprecipitant nucleic acids | GlycoBlue™ | Ambion Applied Biosystems | AM9515-AM9516 |
| Bile acid | Tauroursodeoxycholic Acid, Sodium Salt | EMD Millipore Corp. | 580549 |
| Inducer ER Stress | Tunicamycin | Tocris | 3516 |
| Antibody for IHC | Anti-XBP1 | Abcam | ab109221 |
| Antibody for IHC | Anti-HSPA6/BiP | Abcam | ab21685 |

#### 16S rDNA Illumina sequencing and analysis

Phenol:chloroform-extracted DNAs from stool samples were processed for 16S rDNA Illumina sequencing and analysis.. Primer selection and PCRs were performed as described previously.[1, 2] Briefly, each sample was amplified in triplicate with Golay-barcoded primers specific for the V4 region (F515/R806), combined, and confirmed by gel electrophoresis. PCR reactions contained 18.8 μL RNase/DNase-free water, 2.5 μL 10X High Fidelity PCR Buffer (Invitrogen, 11304–102), 0.5 μL 10 mM dNTPs, 1 μL 50 mM MgSO4, 0.5 μL each of the forward and reverse primers (10 μM final concentration), 0.1 μL Platinum High Fidelity Taq (Invitrogen, 11304–102) and 1.0 μL genomic DNA. Reactions were held at 94°C for 2 min to denature the DNA, with amplification proceeding for 26 cycles at 94°C for 15 s, 50°C for 30 s, and 68°C for 30 s; a final extension of 2 min at 68°C was added to ensure complete amplification. Amplicons were pooled and purified with 0.6x Agencourt AMPure XP beads (Beckman-Coulter, A63882) according to the manufacturer's instructions. The final pooled samples, along with aliquots of the three sequencing primers, were sent to the DNA Sequencing Innovation Lab (Washington University School of Medicine) for sequencing by the 2 X 250bp protocol with the Illumina MiSeq platform.

Read quality control and the resolution of amplicon sequence variants were performed in R version 4.0.5 with DADA2 version 1.18.0 using the Ribosomal Database Project (RDP) trainset 16 release 11.5.[3, 4] Ecological analyses were performed using PhyloSeq and additional R packages in R version 4.3.2.[5] Differentially abundant taxa were identified using DESeqs.[6] 16S sequencing data have been uploaded to the European Nucleotide Archive (accession no. in process). Associated code is available at https://github.com/RachelRodgers/TUDCA.

#### SUPPLEMENTARY FIGURES AND TABLES

| Patient | Adverse Event | Duration (days) | Anatomical location | CTCAE Severity Grade | Is the adverse event serious? | Possibly related to TUDCA | Related to study indication? | Related to concurrent illness? | Related to protocol assessments? | Related to concomitant medication | Related to another cause | Action taken with TUDCA? | Outcome |
|---|---|---|---|---|---|---|---|---|---|---|---|---|---|
| 1 | Dyspepsia | 3 | GI system | 1 | N | Y | N | N | N | N | N | None | Resolved |
| 2 | None | | | | | | | | | | | | |
| 3 | Abdominal cramping | 3 | GI system | 1 | N | Y | N | N | Y (endoscopy) | N | N | None | Resolved |
| 3 | Diarrhea | 3 | GI system | 1 | N | Y | N | N | Y (endoscopy) | N | N | None | Resolved |
| 3 | Dyspepsia | 3 | GI system | 1 | N | Y | N | N | Y (endoscopy) | N | N | None | Resolved |
| 4 | Nausea | 2 | GI system | 1 | N | Y | N | N | N | N | N | None | Resolved |
| 5 | None | | | | | | | | | | | | |
| 6 | None | | | | | | | | | | | | |
| 7 | Dyspepsia | 2 | GI system | 1 | N | Y | N | N | N | N | N | None | Resolved |
| 8 | None | | | | | | | | | | | | |
| 9 | Influenza B | 14 | Respiratory system | 1 | N | N | N | N | N | N | Y | None | Resolved |
| 10 | None | | | | | | | | | | | | |
| 11 | None | | | | | | | | | | | | |
| 12 | Dyspepsia | 5 | GI system | 1 | N | Y | N | N | N | N | N | None | Resolved |
| 12 | Diarrhea | 14 | GI system | 2 | N | Y | Y | N | N | N | N | Dose reduced * | Resolved |
| 14 | Nausea | 85 | GI System | 2 | N | Y | Y | N | N | Y | N | Dose reduced ** | Ongoing at study conclusion |

**Supplementary Table 1. Detailed Safety and Tolerability Reporting**

*Dose reduced to 1000mg/day on 22-Jul-20 and then back to full dose on 30-Jul-20

**Dose reduced to 1000mg/day on 03-May-21 and kept at this dose throughout remainder of the study.

Adverse events categorized according to CTCAE v5.0 criteria and grading.

Grade 1 Mild; asymptomatic or mild symptoms; clinical or diagnostic observations only; intervention not indicated.

Grade 2 Moderate; minimal, local or noninvasive intervention indicated; limiting age appropriate instrumental ADL*.

Grade 3 Severe or medically significant but not immediately life-threatening; hospitalization or prolongation of hospitalization indicated; disabling; limiting self care ADL**.

Grade 4 Life-threatening consequences; urgent intervention indicated.

Grade 5 Death related to AE.

Activities of Daily Living (ADL) Instrumental ADL refer to preparing meals, shopping for groceries or clothes, using the telephone, managing money, etc. Self-care ADL refer to bathing, dressing and undressing, feeding self, using the toilet, taking medications, and not bedridden.

| Laboratory (Avg ± StdDev) | Baseline | Week 2-4 (LFTS only) | Week 6 | P value (paired t-test) |
|---|---|---|---|---|
| WBC | 7.51 ± 2.28 | | 7.32 ± 1.65 | 0.71 |
| HBG | 12.65 ± 1.99 | | 12.39 ± 1.61 | 0.44 |
| PLT | 362.00 ± 107.20 | | 343.69 ± 83.67 | 0.29 |
| Total Protein | 7.42 ± 0.46 | 7.33 ± 0.41 | 7.42 ± 0.42 | 0.96 |
| ALB | 4.20 ± 0.39 | 4.08 ± 0.39 | 4.17 ± 0.36 | 0.77 |
| AST | 20.38 ± 13.81 | 20.30 ± 10.23 | 23.23 ± 10.56 | 0.20 |
| ALT | 20.62 ± 21.41 | 22.00 ± 13.43 | 19.62 ± 10.10 | 0.83 |
| AlkPhos | 82.08 ± 39.27 | 89.70 ± 48.77 | 83.77 ± 41.12 | 0.59 |
| Total Bilirubin | 0.55 ± 0.60 | 0.36 ± 0.11 | 0.52 ± 0.36 | 0.72 |

**Supplementary Table 2. Safety labs.**

| Comparison | Group | Bacteria | Classification | Log2FoldChange | Base Mean | Notes | Reference |
|---|---|---|---|---|---|---|---|
| Baseline vs Post | All Samples | Bacteroidetes | Phylum | 23.3 | 25.20 | Decreased in animal study with TUDCA. Anti-inflammatory. Decreased in patients with CD. | https://www.sciencedirect.com/science/article/pii/S1665268120302143 https://www.jacionline.org/action/showPdf?pii=S0091-6749%2819%2931486-1 https://link.springer.com/article/10.1007/s00535-017-1384-4 |
| Baseline vs Post | All Samples | Firmicutes | Phylum | -24.4 | 33.00 | Increased in animal study with TUDCA. Anti-inflammatory. Decreased in patients with CD. | https://www.sciencedirect.com/science/article/pii/S1665268120302143 https://www.jacionline.org/action/showPdf?pii=S0091-6749%2819%2931486-1 https://link.springer.com/article/10.1007/s00535-017-1384-4 |
| Baseline vs Post | All Samples | Ruminococcus2 | Genus | -22.9 | 10.70 | Increased in healthy controls compared to IBD. | https://www.ncbi.nlm.nih.gov/pmc/articles/PMC7729129/ |
| Baseline vs Post | All Samples | bifidum | Species | -22.5 | 8.00 | Early administration alleviates long term colitis in mice. | https://www.frontiersin.org/journals/microbiology/articles/10.3389/fmicb.2022.916824/full |
| Baseline vs Post | Responders Post (+) and BS (-) | Bacteroidetes | Phylum | 22.8 | 25.20 | Decreased in animal study with TUDCA. Anti-inflammatory. Decreased in patients with CD. | https://www.sciencedirect.com/science/article/pii/S1665268120302143 https://www.jacionline.org/action/showPdf?pii=S0091-6749%2819%2931486-1 https://link.springer.com/article/10.1007/s00535-017-1384-4 |
| Baseline vs Post | Responders Post (+) and BS (-) | dorei | Species | 22.0 | 13.90 | Associated with UC activity. | https://www.ncbi.nlm.nih.gov/pmc/articles/PMC8073534/ |
| Baseline vs Post | Responders Post (+) and BS (-) | Blautia | Genus | -23.5 | 25.00 | Higher abundance in UC patients in remission compared to active UC. | https://www.ncbi.nlm.nih.gov/pmc/articles/PMC8326252/ |
| Baseline vs Post | Responders Post (+) and BS (-) | Firmicutes | Phylum | -23.1 | 19.10 | Increased in animal study with TUDCA. Anti-inflammatory. Decreased in patients with CD. | https://www.sciencedirect.com/science/article/pii/S1665268120302143 https://www.jacionline.org/action/showPdf?pii=S0091-6749%2819%2931486-1 https://link.springer.com/article/10.1007/s00535-017-1384-5 |
| Baseline vs Post | Responders Post (+) and BS (-) | Blautia | Phylum | -23.0 | 17.50 | Higher abundance in UC patients in remission compared to active UC. | https://www.ncbi.nlm.nih.gov/pmc/articles/PMC8326252/ https://www.ncbi.nlm.nih.gov/pmc/articles/PMC7872077/ |
| Baseline vs Post | Responders Post (+) and BS (-) | Ruminococcus2 | Genus | -22.9 | 16.00 | Increased in healthy controls compared to IBD. | https://www.ncbi.nlm.nih.gov/pmc/articles/PMC7729129/ |
| Baseline vs Post | Responders Post (+) and BS (-) | obeum | Species | -22.7 | 14.50 | Aggravates colitis in mice. | https://academic.oup.com/ecco-jcc/article/17/Supplement_1/i1035/7010350 |
| Baseline | Responders (+) and Non-Responders (-) | Dorea | Genus | 24.8 | 290.80 | Association with mucosal healing and increased Dorea genus. Increased abundance in CD compared to healthy subjects. | https://www.ncbi.nlm.nih.gov/pmc/articles/PMC7511440/ |
| Baseline | Responders (+) and Non-Responders (-) | lactatifermentans | Species | 23.1 | 89.30 | No known association with inflammatory bowel disease. | |
| Baseline | Responders (+) and Non-Responders (-) | Firmicutes | Phylum | 22.4 | 53.00 | Increased in animal study with TUDCA. Anti-inflammatory. Decreased in patients with CD. | https://www.sciencedirect.com/science/article/pii/S1665268120302143 https://www.jacionline.org/action/showPdf?pii=S0091-6749%2819%2931486-1 https://link.springer.com/article/10.1007/s00535-017-1384-4 |
| Baseline | Responders (+) and Non-Responders (-) | Firmicutes | Phylum | 21.3 | 23.50 | Increased in animal study with TUDCA. Anti-inflammatory. Decreased in patients with CD. | https://www.sciencedirect.com/science/article/pii/S1665268120302143 https://www.jacionline.org/action/showPdf?pii=S0091-6749%2819%2931486-1 https://link.springer.com/article/10.1007/s00535-017-1384-4 |
| Baseline | Responders (+) and Non-Responders (-) | Ruminococcus2 | Genus | 21.1 | 21.10 | Increased in healthy controls compared to IBD. | https://www.ncbi.nlm.nih.gov/pmc/articles/PMC7729129/ |
| Baseline | Responders (+) and Non-Responders (-) | uniformis | Species | 20.9 | 17.90 | Significantly higher in healthy controls compared to patients with UC. | https://pubmed.ncbi.nlm.nih.gov/37834117/#:~:text=Previous%20studies%20have%20demonstrated%20that,UC%20has%20not%20been%20characterized. |
| Baseline | Responders (+) and Non-Responders (-) | bifidum | Species | 20.9 | 20.40 | Early administration alleviates long term colitis in mice. | https://www.frontiersin.org/journals/microbiology/articles/10.3389/fmicb.2022.916824/full |
| Baseline | Responders (+) and Non-Responders (-) | Blautia | Genus | 20.6 | 23.00 | Higher abundance in UC patients in remission compared to active UC. | https://www.ncbi.nlm.nih.gov/pmc/articles/PMC8326252/ https://www.ncbi.nlm.nih.gov/pmc/articles/PMC7872077/ |
| Baseline | Responders (+) and Non-Responders (-) | Veillonella | Genus | 20.4 | 12.20 | Higher levels in recently diagnosed UC. | https://link.springer.com/article/10.1007/s10238-022-00917-x#:~:text=The%20gut%20microbiome%20of%20recently,to%20predict%20response%20to%20therapy. |
| Baseline | Responders (+) and Non-Responders (-) | Bifidobacterium | Genus | 11.5 | 359.80 | Increased abundance in patients with IBD compared to controls. Some evidence that decrease in bifidobacterium abundance increases TUDCA accumulation by reducing bile bile salt hydrolase production. | https://www.ncbi.nlm.nih.gov/pmc/articles/PMC3911339/ |
| Post | Responders (+) and Non-Responders (-) | Bifidobacterium | Genus | 26.2 | 765.90 | Increased abdunance in patients with IBD compared to controls. Some evidence that decrease in bifidobacterium abundance increases TUDCA accumulation by reducing bile bile salt hydrolase production. | https://www.ncbi.nlm.nih.gov/pmc/articles/PMC3911339/ |
| Post | Responders (+) and Non-Responders (-) | Clostridium_IV | Genus | 23.6 | 121.90 | Significant role in maintaining intestinal function by producing butyrate. Reduced in occurrence of UC. | https://www.ncbi.nlm.nih.gov/pmc/articles/PMC5712343/#:~:text=Clostridium%20clusters%20IV%20and%20XIVa%20were%20demonstrated%20to%20play%20a,UC%20(43%2C%2044). |
| Post | Responders (+) and Non-Responders (-) | Clostridium_III | Genus | 21.7 | 29.60 | No known association with inflammatory bowel disease. | |
| Post | Responders (+) and Non-Responders (-) | Bacteroides | Genus | 21.4 | 23.80 | Lower levels associated with active UC. | https://pubmed.ncbi.nlm.nih.gov/27999802/ |
| Post | Responders (+) and Non-Responders (-) | bifidum | Species | 21.1 | 18.60 | Early administration alleviates long term colitis in mice. | https://www.frontiersin.org/journals/microbiology/articles/10.3389/fmicb.2022.916824/full |
| Post | Responders (+) and Non-Responders (-) | Ruminococcus2 | Genus | 20.5 | 12.50 | Increased in healthy controls compared to IBD. | https://www.ncbi.nlm.nih.gov/pmc/articles/PMC7729129/ |
| Post | Responders (+) and Non-Responders (-) | Clostridium_XIVa | Genus | -25.0 | 50.50 | Reduced in the occurrence of UC. | https://www.ncbi.nlm.nih.gov/pmc/articles/PMC5712343/#:~:text=Clostridium%20clusters%20IV%20and%20XIVa%20were%20demonstrated%20to%20play%20a,UC%20(43%2C%2044). |

**Supplementary Table 3. Microbiome Bacteria Specifics.**

Further details regarding bacteria noted in Figure 6C and Supplementary Figure 7. Table includes corresponding comparison and group, bacteria name, classification, log2 fold change, base mean, relevant notes, and links to references.

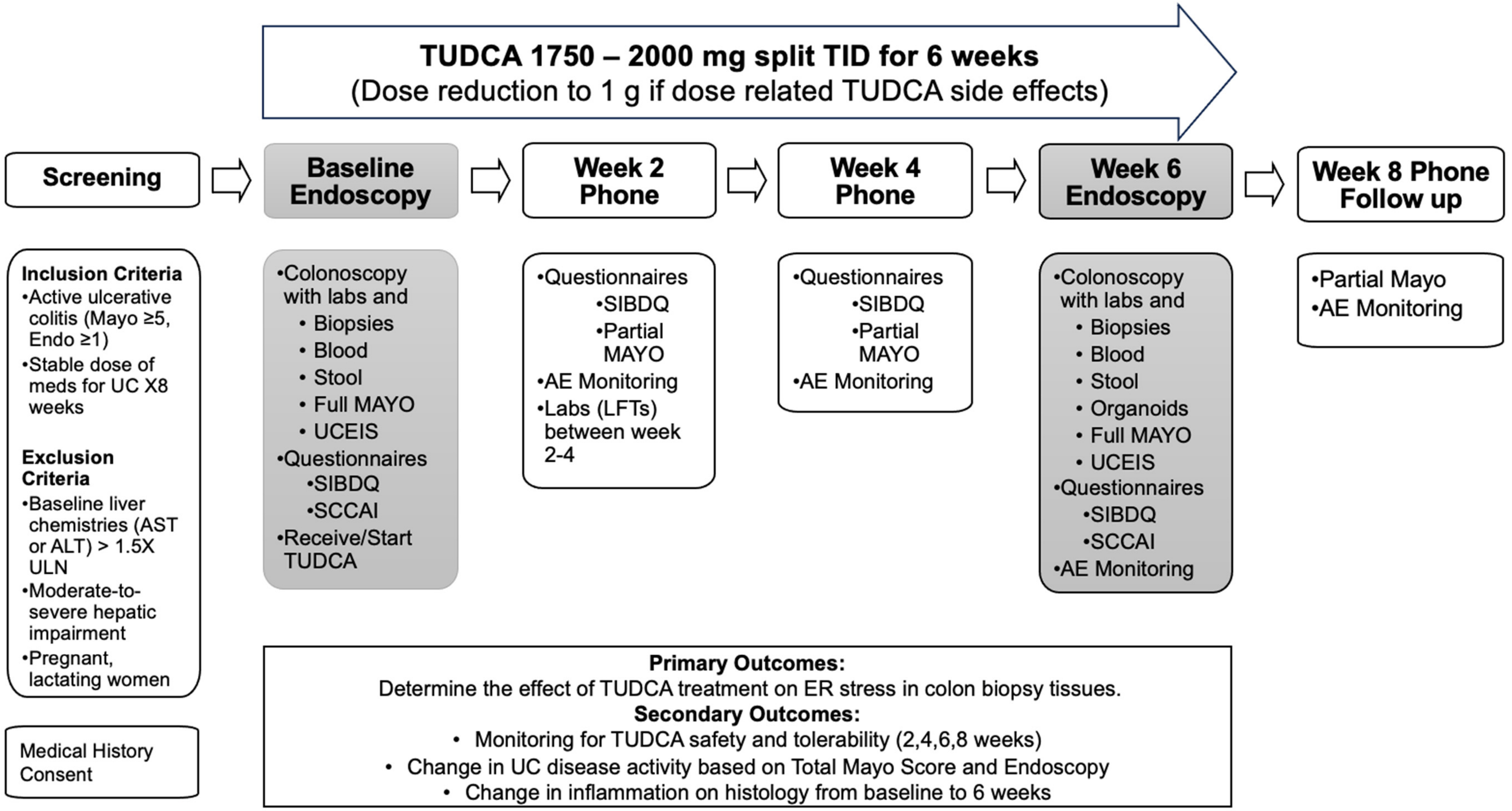


**Supplementary Figure 1. Clinical Trial Schema**

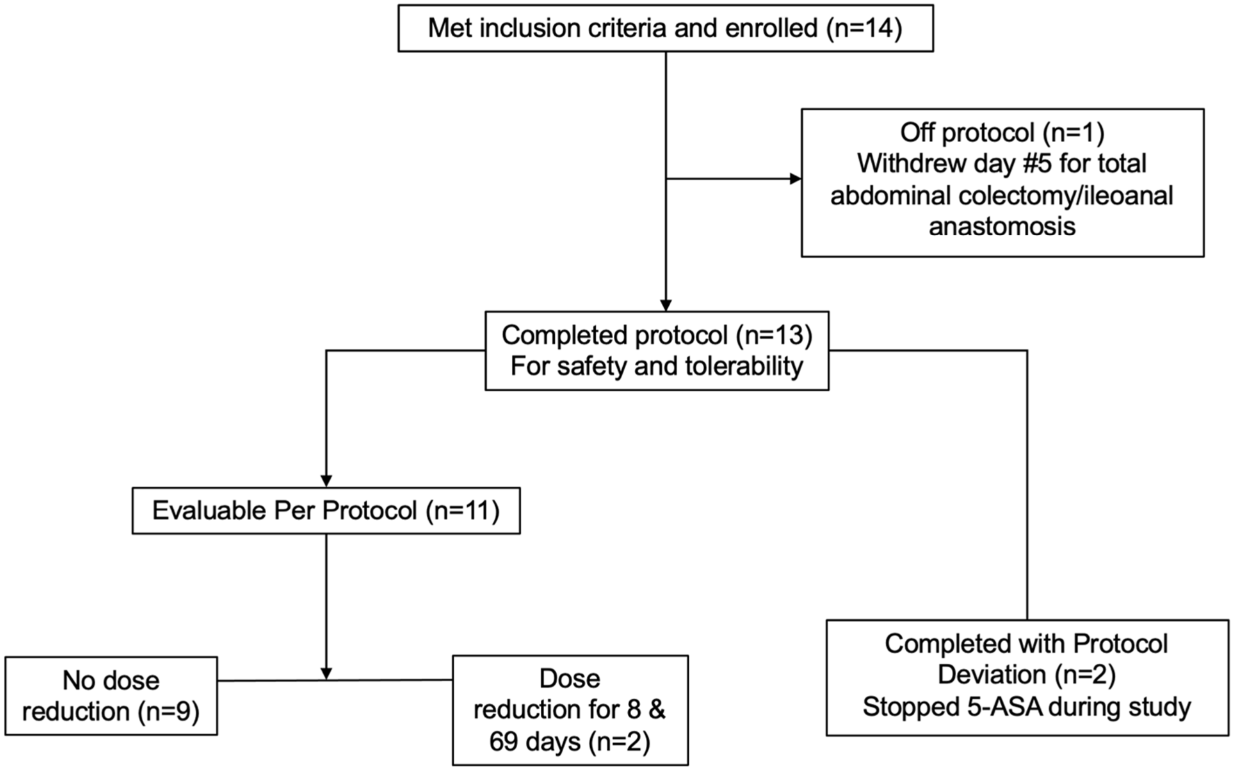


**Supplementary Figure 2. Flowchart of Study Patients**

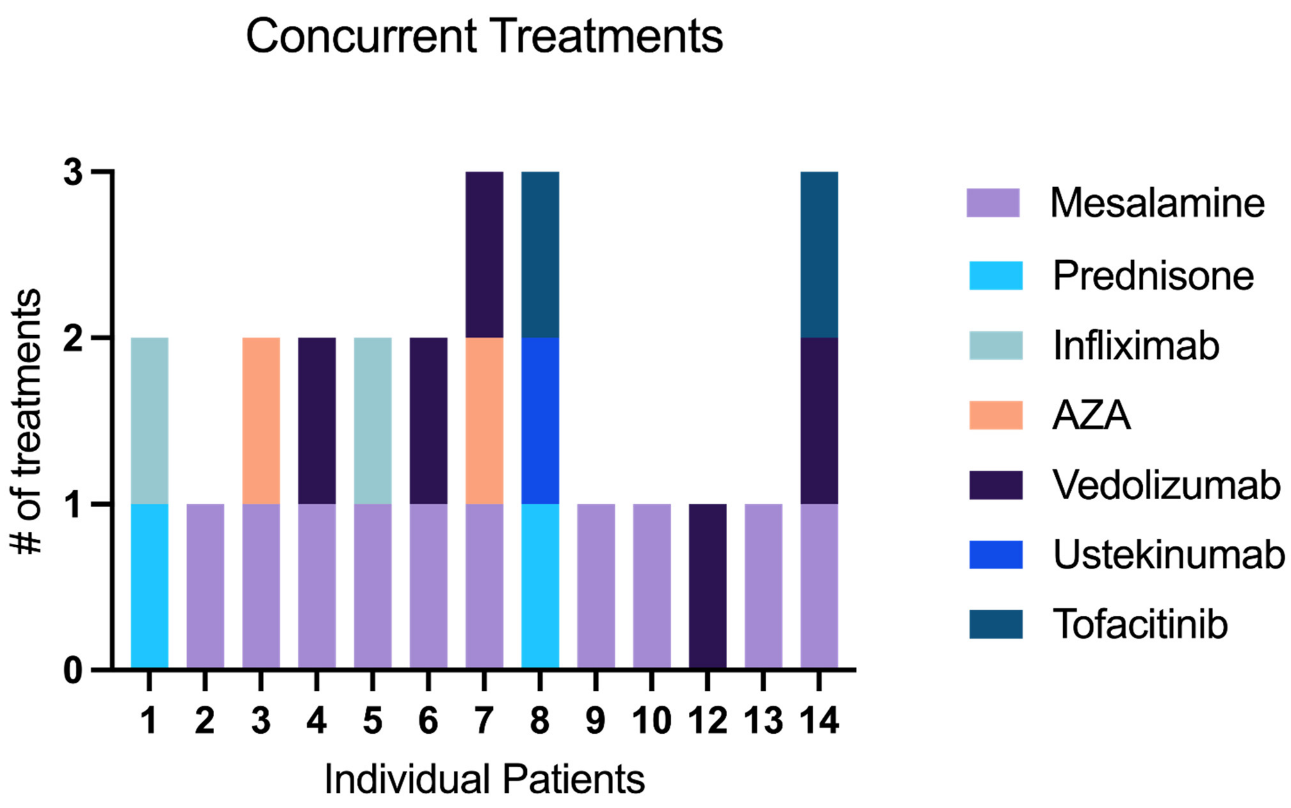


**Supplementary Figure 3. Concurrent treatments for individual patients**

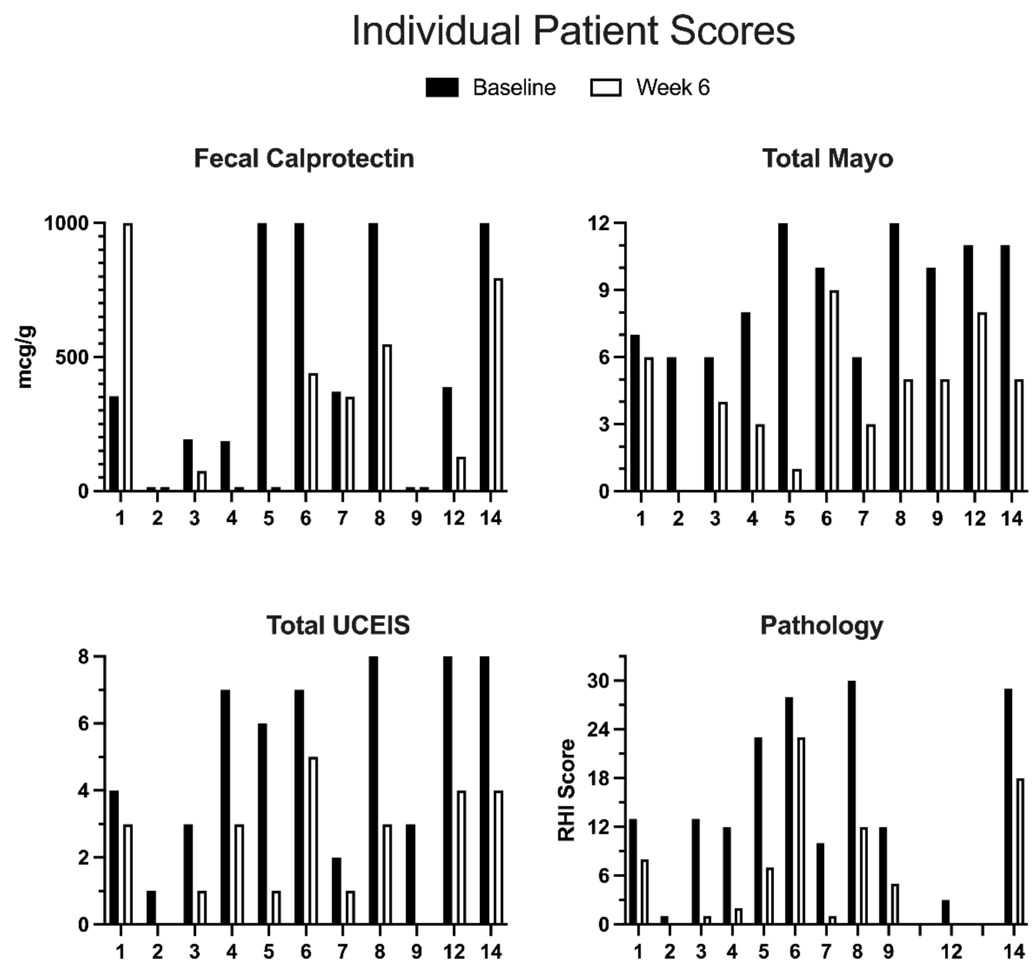


**Supplementary Figure 4. Changes in disease activity by individual patient**

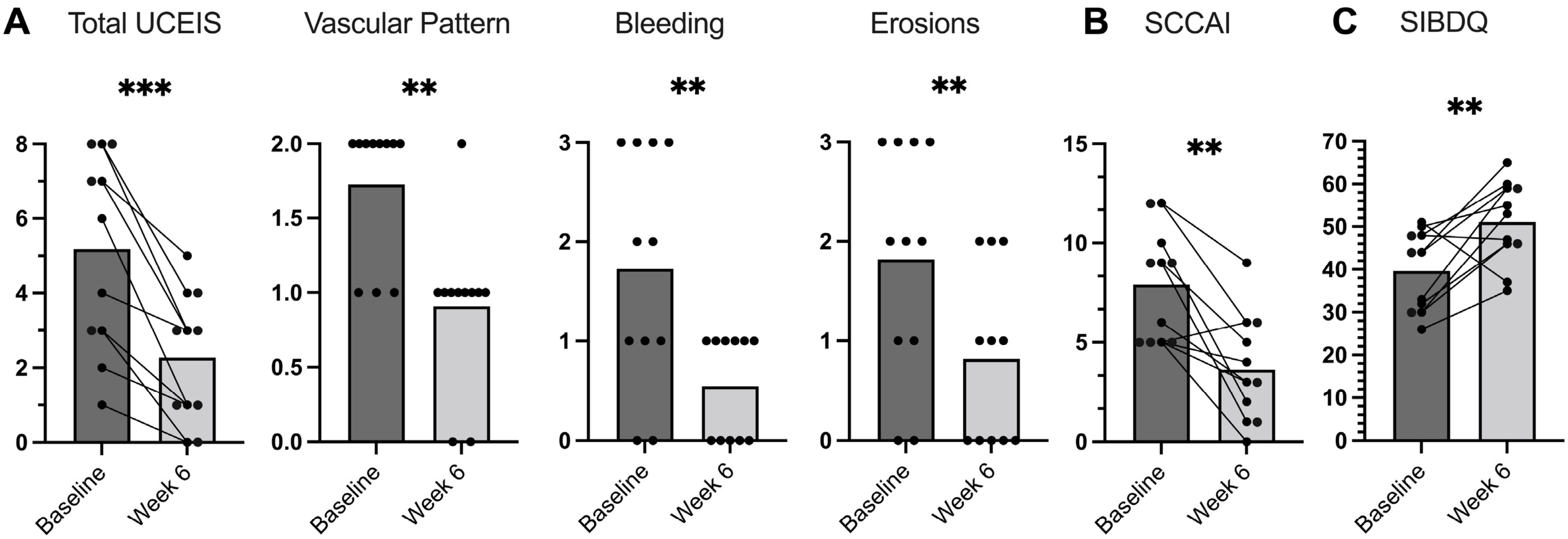


**Supplementary Figure 5. Additional measures of endoscopic and clinical disease activity before and after TUDCA. A)** The total and subscores for the Ulcerative Colitis Endoscopic Index of Severity (UCEIS). **B)** Simple Clinical Colitis Activity Index (SCCAI). **C)** Short Inflammatory Bowel Diseases Questionnaire (SIBDQ). Statistics by two-tailed Student's t test (total UCEIS) and two-tailed Wilcoxon matched-pairs signed rank test (UCEIS subscores, SCCAI, and SIBDQ). P values *<.05, **<.01, ***<.001.

**A.**

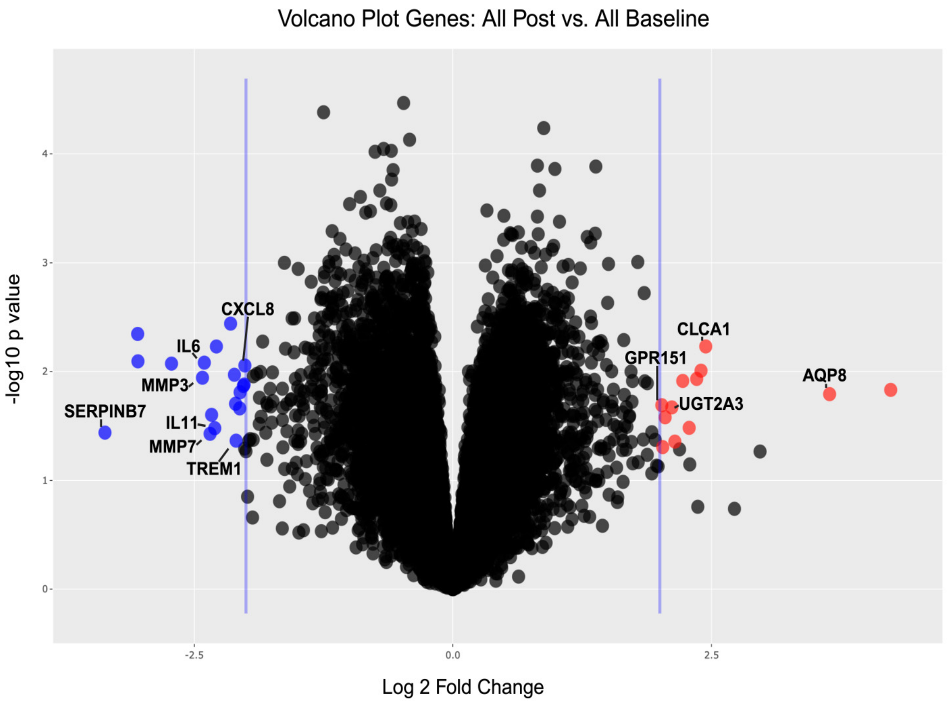


**B.**

| Ensembl ID | Gene Name | logFC | P value | Gene Expression | Comments | Reference |
|---|---|---|---|---|---|---|
| ENSG00000166396 | SERPINB7 | -3.36 | 0.04 | Down | Upregulated in UC | PMID: 35719390 |
| ENSG00000136244 | IL6 | -2.40 | 0.01 | Down | Upregulated in IBD | PMID: 21848856 |
| ENSG00000149968 | MMP3 | -2.42 | 0.01 | Down | Increased in IBD | PMID: 25611261 |
| ENSG00000095752 | IL11 | -2.30 | 0.03 | Down | Associated with IBD | PMID: 31986085 |
| ENSG00000137673 | MMP7 | -2.35 | 0.04 | Down | Associated with UC | PMID: 36275703 |
| ENSG00000169429 | CXCL8 | -2.01 | 0.01 | Down | Associated with UC | PMID: 33706134 |
| ENSG00000124731 | TREM1 | -2.10 | 0.04 | Down | Correlates with gene activity in UC | PMID: 33537747 |
| ENSG00000103375 | AQP8 | 3.64 | 0.02 | Up | UC associated with decrease in AQP8 | PMID: 23595390 |
| ENSG00000016490 | CLCA1 | 2.44 | 0.01 | Up | Associated with goblet cell mucus production | PMID: 30914450 |
| ENSG00000135220 | UGT2A3 | 2.12 | 0.02 | Up | Role in bile acid metabolism | PMID: 23756265 |
| ENSG00000173250 | GPR151 | 2.02 | 0.02 | Up | Regulates glucose metabolism | PMID: 36456565 |

**Supplementary Figure 6. Volcano plot of gene expression changes after TUDCA treatment. A.** Colored genes met the criteria of >2.0 (RED) or <-2.0 (BLUE) fold change and P<0.05. Genes related to UC are labeled. B. Table includes Ensembl ID, gene name, log fold change, p value, associated gene expression and the PMID of the associated reference.

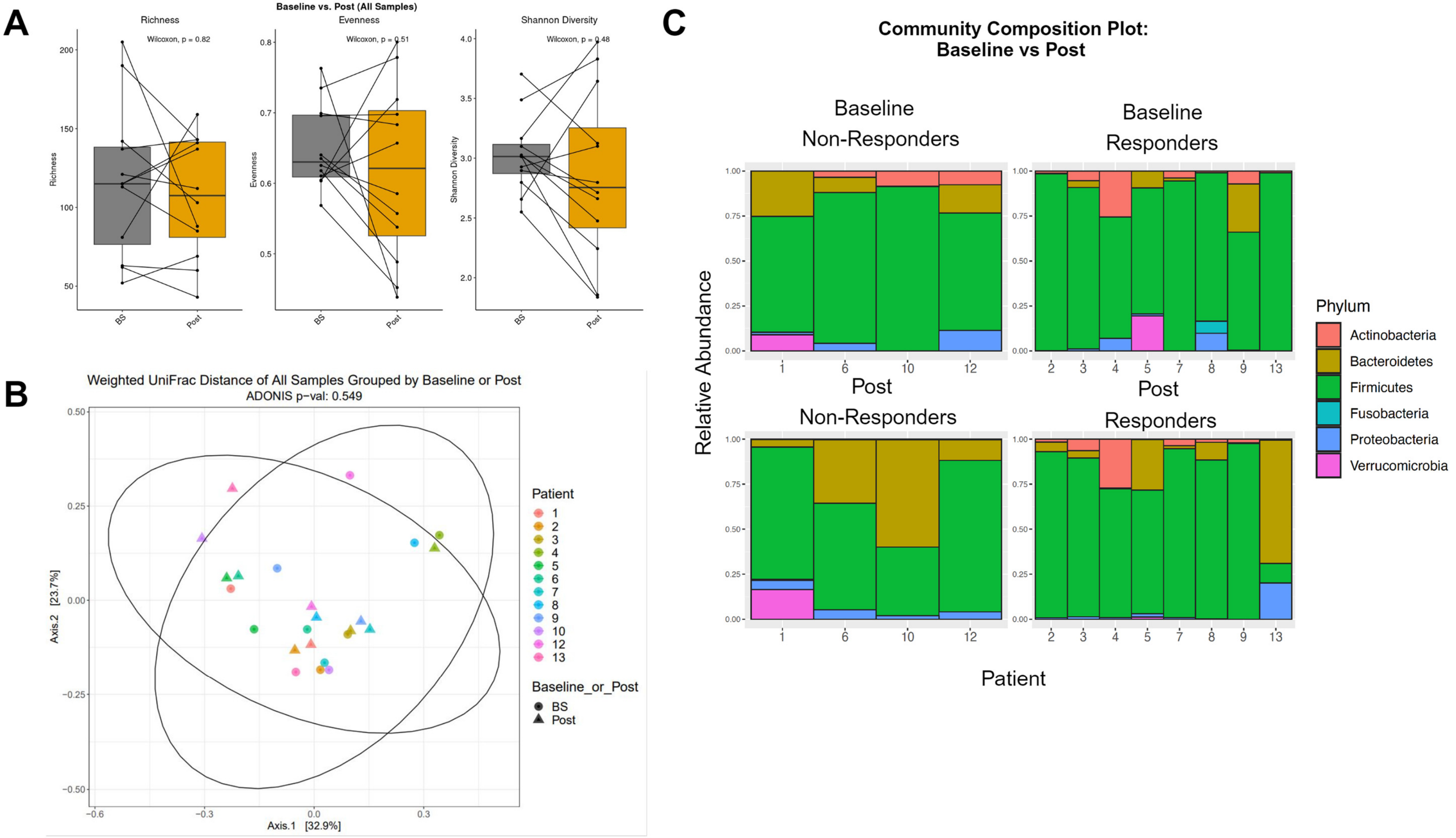


**Supplementary Figure 7.** Microbiome diversity and composition changes with TUDCA treatment. Stools from all patients with pre and post treatment samples were analyzed by 16S. **A)** Alpha diversity for all baseline (BS) and post TUDCA treatment samples. **B)** Beta diversity from BS and post treatment **C)** Community composition of bacteria further grouped by responders (clinical and mucosal healing) or non-responders.

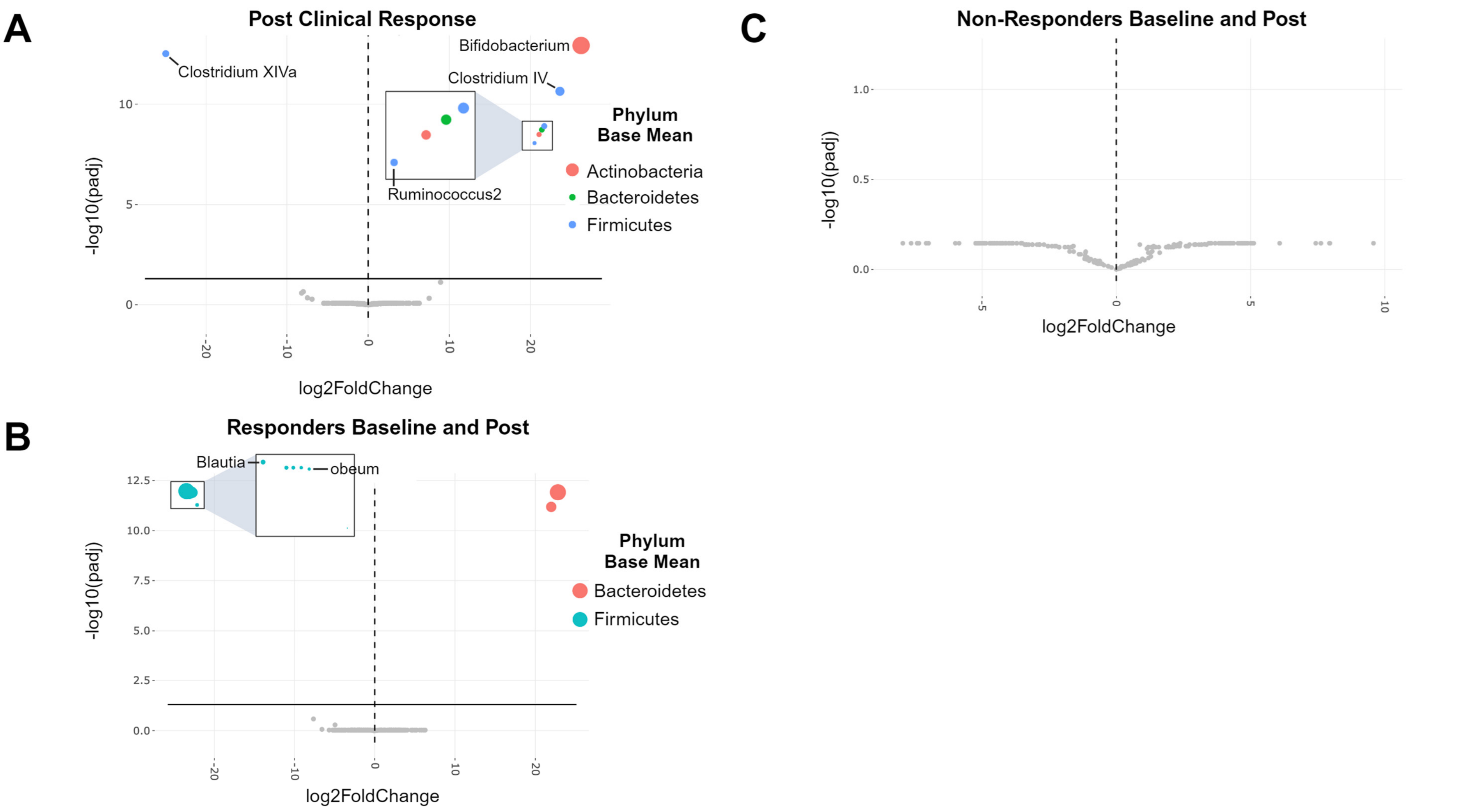


**Supplementary Figure 8. Stool biomarker analysis**. Taxonomic assignment of the 16S data was made using DADA2 with the RDP v16 training set. Differentially abundant taxa were identified using DESeqs. **A)** All post TUDCA treatment stools grouped by responders (right of midline) vs. non-responders (left of midline). **B)** All responders baseline (left) vs post treatment (right) samples. **C)** All non-responders grouped by baseline (left) vs post treatment (right). All circles in color and above solid horizontal black line represent those with significant ($p<0.05$) differential expression.
